# Immune Memory Response After a Booster Injection of mRNA-1273 for Severe Acute Respiratory Syndrome Coronavirus-2 (SARS-CoV-2)

**DOI:** 10.1101/2021.09.29.21264089

**Authors:** Laurence Chu, David Montefiori, Wenmei Huang, Biliana Nestorova, Ying Chang, Andrea Carfi, Darin K. Edwards, Judy Oestreicher, Holly Legault, Bethany Girard, Rolando Pajon, Jacqueline M. Miller, Rituparna Das, Brett Leav, Roderick McPhee

## Abstract

Rising breakthrough infections of coronavirus-2 (SARS-CoV-2) in previously immunized individuals has raised concerns for a booster to combat suspected waning immunity and new variants. Participants immunized 6-8 months earlier with a primary series of two doses of 50 or 100 µg of mRNA-1273 were administered a booster injection of 50 µg of mRNA-1273. Neutralizing antibody levels against wild-type virus and the Delta variant at one month after the booster were 1.7-fold and 2.1-fold higher, respectively, than those 28 days post primary series second injection indicating an immune memory response. The reactogenicity after the booster dose was similar to that after the second dose in the primary series of two doses of mRNA-1273 (50 or 100 µg) with no serious adverse events reported in the one-month follow-up period. These results demonstrate that a booster injection of mRNA-1273 in previously immunized individuals stimulated an immune response greater than the primary vaccination series.

## Introduction

Several vaccines for severe acute respiratory coronavirus-2 (SARS-CoV-2) have been authorized for emergency use, and one vaccine has been approved by the U.S. FDA for immunization of individuals ≥16 years of age. On September 22, 2021, the U.S. FDA approved a third dose of BNT162b2 in individuals ≥65 years of age, those 18 to 64 years of age at high risk of severe COVID-19, and those who have institutional or occupational exposure to SARS-CoV-2 which puts them at high risk of serious complications of COVID-19.^1^ SARS-CoV-2 vaccines have been administered worldwide to billions of people.^2–4^ These vaccines are very efficacious in preventing hospitalizations and deaths due to COVID-19.^5–7^ mRNA-1273 is a lipid nanoparticle-encapsulated messenger RNA encoding the S protein of the Wuhan-Hu-1 isolate with 2 proline mutations introduced to stabilize the S protein into the prefusion conformation. mRNA-1273 has demonstrated immune responses against SARS-CoV-2, efficacy against COVID-19 disease in adults and adolescents and an acceptable safety and tolerability profile in several clinical trials.^5, 8–11^ Based on a vaccine efficacy of 94.1% against COVID-19 after a median follow up of 64 days^5^, demonstrated in the phase 3 COVE study, mRNA-1273 received emergency use authorization from the Food and Drug Administration in December, 2020 for use in adults 18 years of age or older.^12^ Recently, the final efficacy analysis of the blinded part of the study demonstrated a vaccine efficacy of 93.2% over 5.3 months of follow-up.^13^

There are concerns that the efficacy of the SARS-CoV-2 vaccines may decrease in the future due to waning immunity and/or emerging viral variants. Variants of SARS-CoV-2 with amino acid changes in the spike protein (S) and elsewhere in the viral genome are circulating around the world.^14^ Variants such as Alpha (B.1.1.7), Beta (B.1.351), Gamma (P.1), Delta (B.1.617.2), Epsilon (B.1.427; B.1.429), Iota (B.1.526) and Mu (B.1.621) are highly transmissible ^14^ and have become the predominant cause of SARS-CoV-2 outbreaks in large parts of the world (e.g., B.1.617.2 [Delta] in the United States, Europe, India, and South Africa; P.1 [Gamma] in Brazil and B.1.621 [Mu] in Colombia).^15, 16^ Although the overall impact of variants on vaccine efficacy remains to be determined, some of these variants have decreased susceptibilities to neutralizing antibodies induced by current vaccines.^17–23^ Furthermore, in a small study, immunity against variants wanes in vaccinated individuals such that approximately one-half did not have detectable neutralizing antibodies to the Delta variant 6 months after vaccination with mRNA-1273.^23^

Although mRNA vaccine effectiveness against severe disease and hospitalization due to SARS-CoV-2 has remained high, in a large observational study performed by the Mayo Clinic, the mRNA vaccines were less effective against SARS-CoV-2 infections at a time when the Delta variant was prevalent.^24^ Similar declines in mRNA vaccine effectiveness against SARS-CoV-2 infection were observed in nursing home residents in the time period of June 21 to August 1, 2021 when the Delta variant was prevalent, in frontline healthcare workers, and in a real-world effectiveness study in Qatar.^25–27^

These findings with variants and waning immunity after two doses of mRNA vaccines suggest that a booster vaccine injection may be beneficial. In the blinded portion (Part A) of this phase 2 trial of mRNA-1273, participants received two injections of placebo, 50 µg or 100 µg of mRNA-1273. Preliminary results of Part A demonstrated robust immune responses through 1 month after the second injection of mRNA-1273 and an acceptable safety profile in healthy adults aged 18 years and older.^11^ In Part B, the Open-label Interventional phase of this study, participants who received a 2-dose (50 µg or 100 µg of mRNA-1273) prime series in Part A, were offered a single booster dose of mRNA-1273 (50 μg). Here we report the immunogenicity, safety and reactogenicity after the booster dose, as well as antibody persistence before the booster dose (approximately 209 days after the primary series).

## Methods

### Study design

This phase 2 study (NCT04405076) enrolled 600 participants to receive placebo, or 50 or 100 µg of mRNA-1273 (randomized 1:1:1; Figure 1A) in two cohorts of participants ≥18 to <55 years old (Cohort 1) and participants ≥ 55 years old (Cohort 2) in the observer-blind and placebo-controlled part of the study (Part A; Figure 1).

**Figure 1.**
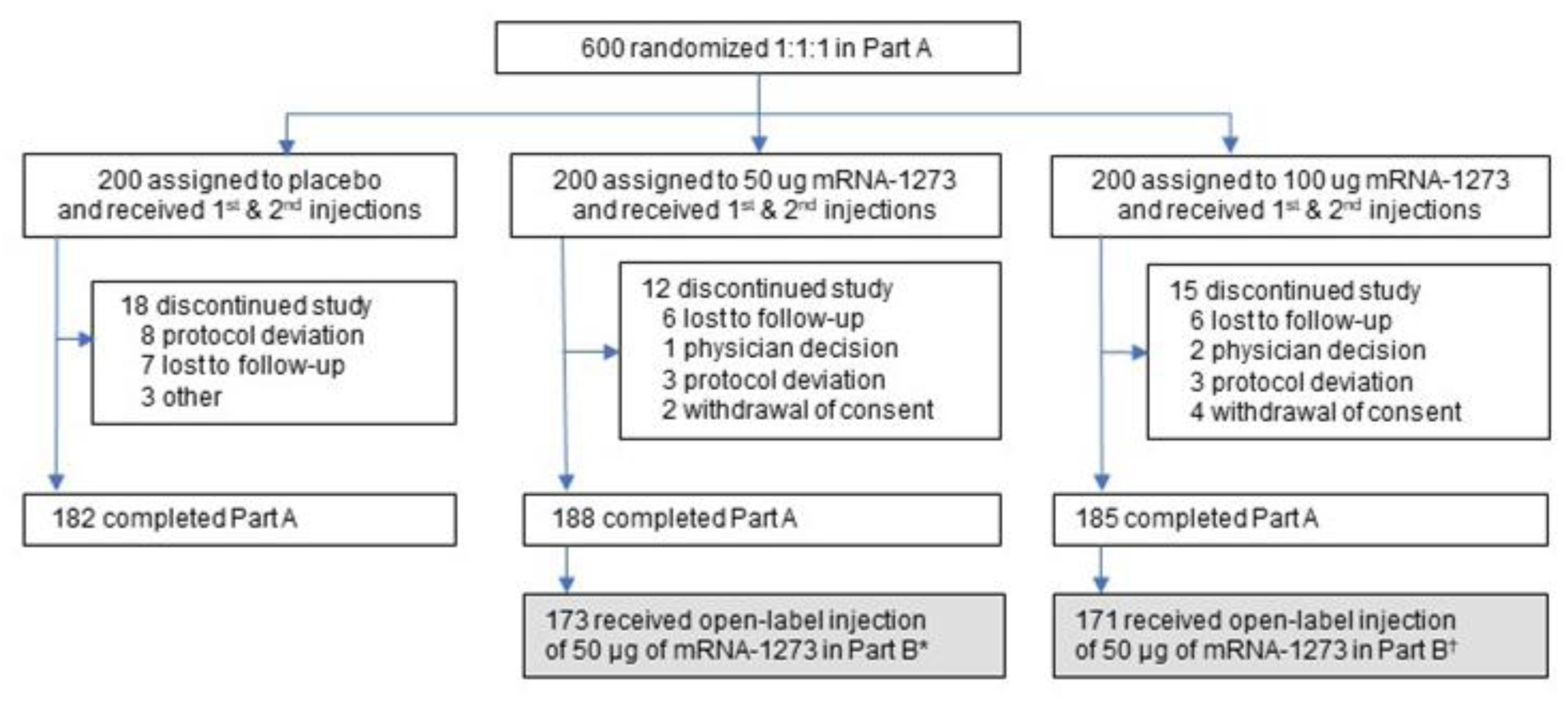
Trial Profile for Phase 2, Parts A and B. Participants who received two doses of mRNA-1273 in Part A were offered a booster injection of 50 µg of mRNA-1273 in Part B. Completion of Part A was defined as participants who completed 6 months of follow up after the last injection received in Part B (open-label phase). Data cut-off June 11, 2021. *15 participants declined to receive a booster of 50 μg mRNA-1273. ^†^14 participants declined to receive a booster of 50 μg mRNA-1273.

Preliminary immunogenicity and safety results were previously reported.^11^ Previously, the primary efficacy endpoint for mRNA-1273 against COVID-19 was met in the phase 3 efficacy study (COVE study)^5^, 554 participants were unblinded in Part B at least 5.9 months or more after enrollment in Part A of the study. In total, 173 or 171 participants who had initially received 2 injections of 50 µg or 100 µg of mRNA-1273, respectively, then received a single booster of 50 µg of mRNA-1273.

### Trial participants

Eligible participants in Part A were male or female, 18 years of age or older at screening and in good general health according to the investigator. For Part B, participants must have been previously enrolled in the mRNA-1273 phase 2 study. Exclusion criteria were pregnancy or breastfeeding, acute illness or febrile (body temperature ≥ 38.0°C/100.4°F) 24 hours prior to or at screening, or current treatment with investigational agents for prophylaxis against COVID-19.

### Randomization and Blinding

There were two age cohorts in this phase 2 study: participants ≥18 to <55 years old in Cohort 1 and participants ≥ 55 years old in Cohort 2. Within each age cohort, approximately 300 participants were randomized in 1:1:1 ratio to receive 50 µg of mRNA-1273, 100 µg of mRNA-1273, or placebo in Part A. The randomization was performed in a blinded manner using a centralized Interactive Response Technology. Vaccine dose preparation and administration during Part A were performed by unblinded pharmacy personnel who did not participate in any other aspects of the study. A limited number of the sponsor team and clinical research organization (CRO) were unblinded to enable the primary analysis at one month after the second dose of mRNA-1273 in Part A. All study staff, participants, CRO and sponsor personnel remained blinded to dosing assignment until the study was unblinded, upon implementation of Part B of the study, following Emergency Use Authorization of mRNA-1273 in the United States.

Participants with negative baseline SARS-CoV-2 status (n=1080) were randomly selected from the phase 3 COVE trial participants in the mRNA-1273 group to form an Immunogenicity Subset which was subsequently used as the historical comparator arm for the phase 2 Part B immunobridging analysis. Of the 1080 selected participants from the phase 3 COVE trial mRNA-1273 group, 25 were further excluded from the Per Protocol Immunogenicity Subset for the following reasons: had HIV infection (18 participants), received dose 2 outside of the pre-specified window (5 participants), did not receive dose 2 per schedule (1 participant), or had major protocol deviations (1 participant). Thus, 1055 participants were included in the Per Protocol Immunogenicity Subset from the phase 3 COVE trial.

### Trial vaccine

The mRNA-1273 vaccine is a lipid nanoparticle containing an mRNA that encodes the SARS-CoV-2 spike glycoprotein of the Wuhan-HU-1 isolate.^9, 11^ The placebo and the mRNA-1273 vaccine were administered in the deltoid as an intramuscular injection according to a 2-dose regimen in Part A with the first dose given on day 1 and the second on day 29 (28 days after dose 1). In Part B, 50 µg of mRNA-1273 was administered intramuscularly in the deltoid as a single booster injection at Open-label Day 1 (OL-D1) to the treatment groups originally vaccinated with either dose regimen of mRNA-1273. The volume administered in both Part A and Part B in each injection was 0.5 mL containing 50 or 100 µg of mRNA-1273, or saline (placebo).

### Study outcomes

Details regarding the design of Part A of the study have been previously published.^28^ The primary safety objective of Part B was to evaluate the safety and reactogenicity of 50 µg of mRNA-1273 administered as a single booster dose 6 months or more after a priming series of 50 µg or 100 µg of mRNA 1273. The primary safety endpoints were solicited local and systemic adverse reactions (ARs) through 7 days after each injection, unsolicited treatment-emergent adverse events (TEAEs) through 28 days after each injection, medically-attended AEs (MAAEs) and serious AEs (SAEs) throughout the entire study period.

The primary immunogenicity objective was to evaluate the immunogenicity of 50 µg of mRNA-1273 administered as a single booster dose administered at least 6 months after a two-dose priming series with 50 or 100 µg of mRNA-1273 as compared to 100 µg of mRNA-1273 administered as 2 doses 28 days apart in the pivotal phase 3 efficacy and safety study (COVE-P301), as assessed by the level of SARS-CoV-2-specific neutralizing antibody (nAb). The coprimary endpoints for noninferiority were: (i) Geometric mean (GM) titers of serum nAb and (ii) Seroresponse rates for nAb based on the pseudovirus neutralizing antibody assay. The secondary immunogenicity objective was to evaluate the immunogenicity of 50 µg of mRNA-1273 vaccine administered as a single booster dose as assessed by the titers of bAb. Levels of SARS-CoV-2-specific bAb were measured by enzyme-linked immunosorbent assay (ELISA) and a SARS-CoV-2 Meso-Scale Discovery (MSD) 3-PLEX assay on OL-D1 (pre-boost) and OL-D29 (28 days after the booster injection). Seroresponse was defined in 3 ways: i) seroresponse (specific to the ID50 titer in the pseudovirus neutralizing antibody assay) was defined as a change from below the lower limit of quantification (LLOQ) at pre-booster (or pre-dose 1) to equal or above LLOQ at 28 days after the booster (or 28 days after the primary series), or at least a 3.3-fold rise at 28 days after the booster (or 28 days after the primary series) if pre-booster (or pre-dose 1) titer was equal to or above LLOQ; ii) Seroresponse (4-fold rise) was defined as a change of titer from below the LLOQ at pre-booster (or pre-dose 1) to equal to or above 4 × LLOQ at 28 days after the booster (or 28 days after the primary series), or a 4-times or higher ratio in participants with titers above the LLOQ at pre-booster (or pre-dose 1); and iii) Seroresponse (4-fold rise from baseline) was defined as a change of titer from below the LLOQ at pre-dose 1 to equal to or above 4 × LLOQ at 28 days after the booster (or 28 days after the primary series), or a 4-times or higher ratio in participants with titers above the LLOQ at pre-dose 1. Definition iii) was applied on participants in the Phase 2 study only, primary series in Part A and booster in Part B.

### Safety assessment

Solicited local and systemic adverse reactions were recorded daily by participants in an electronic diary during the 7 days after vaccine administration. Any solicited adverse reaction that persisted beyond Day 7 was reported in the electronic diary until resolution. Oral body temperatures were measured daily. If applicable, the size of injection site erythema and swelling/induration were measured and recorded. In Part B, trained site personnel called trial participants every 4 weeks to assess safety beginning 3 months after the booster dose.

### Immunogenicity assessments

#### SARS-CoV-2 Spike-Pseudotyped Virus Neutralization Assay

This validated assay quantifies SARS-CoV-2 neutralizing antibodies by using lentivirus particles that express SARS-CoV-2 spike proteins (Wuhan-Hu-1 isolate including D614G or the Delta variant [B.1.617.2 AY.3; Wuhan-Hu-1 isolate containing spike mutations T19R, G142D, Δ156-157, R158G, L452R, T478K, D614G, P681R, D950N]) on their surface and contain a firefly luciferase reporter gene for quantitative measurements of infection by relative luminescence units (RLU).^29^ The virus was applied to transduced 293T cells expressing high levels of ACE2 (293T/ACE2 cells), with or without pre-incubation with antibodies (control antibodies or serum samples); the presence of neutralizing antibodies reduced infection and resulted in lower RLUs. Serial dilution of antibodies or serum samples were used to produce a dose-response curve. Neutralization was measured as the serum dilution at which the RLU was reduced by 50% (50 percent inhibitory dose [ID50]) or 80% (80 percent inhibitory dose [ID80]) relative to mean RLU in virus control wells (cells + virus but no control antibody or sample) after subtraction of the mean RLU in cell control wells (cells only).

#### SARS-CoV-2 Meso-Scale Discovery (MSD) 3-PLEX assay

This quantitative electrochemiluminescence (ECL) method is an indirect binding ECL method designed to detect SARS-CoV-2 antibodies [SARS-CoV-2 Spike (S; Wuhan-Hu-1 isolate including D614G), nucleocapsid (N), and receptor binding domain (RBD) antibodies] in human serum. The assay is based on the Meso-Scale Discovery (MSD) technology which employs capture molecule MULTI-SPOT® microtiter plates fitted with a series of electrodes. Using an MSD MESO Sector S 600 detection system, an electrical current was applied to the custom microtiter plates leading to a light emission by SULFO-TAGTM through a series of oxidation-reduction reactions involving ruthenium and tripropylamine (TPA). A plate reader measured the intensity of emitted light to provide quantitative measures of analytes in samples.

For this bioassay, a 10-spot custom SARS-CoV-2 3-PLEX plate coated with SARS-CoV-2 antigens (S [containing D614G], N, and RBD) was used. Anti-SARS-CoV-2 antibodies present in the test sample bound to the antigen coated plates and formed an antibody-antigen complexes. These complexes were detected by adding SULFO-TAGTM-labeled antibodies, which bind to the antibody-antigen complexes. Addition of TPA in a buffer solution resulted in ECL that was measured in relative light units (RLU) using the MSD SECTOR S 600 Plate Reader. Antibody concentrations were determined by interpolating their ECL response using the standard curve generated from a serially diluted reference standard.

#### SARS-CoV-2 S-2P IgG ELISA

This validated assay uses microtiter plates coated with commercially available SARS-CoV-2 full-length spike glycoprotein [Wuhan-Hu-1 isolate including D614G]. Serum containing the SARS-CoV-2 IgG antibody was added to the plates. Bound antigen-antibody complex was detected using purified goat anti-human IgG horseradish peroxidase conjugate. Color development occurred with the addition of 3,3′,5,5′tetramethylbenzidine substrate and color intensity was measured spectrophotometrically (450 nm). The intensity of the color was directly proportional to the IgG antibody concentration. Quantitation of the human IgG antibody to SARS-CoV-2, or antibody concentration (AU/mL), was determined by interpolation from a standard curve analyzed on each assay plate.

### Statistical analyses

The results for the two groups that received a booster injection after a primary series of two doses of 50 µg or 100 µg of mRNA-1273 were expected to be similar and have been combined for the immunogenicity analysis to increase the statistical power for comparisons to the historical control from the phase 3 COVE trial.

The safety analyses were descriptive without pre-specified statistical criteria. The Safety Set for Part B booster included all participants who were randomized in Part A and received a booster injection during Part B. The Solicited Safety Set for Part B booster consisted of all participants who were randomized to mRNA-1273 (50 or 100 µg) in Part A, received a booster injection during Part B, and contributed any solicited adverse reaction data (i.e., reported at least one post-baseline solicited safety assessment in Part B). The Solicited Safety Set was used for the analyses of solicited ARs. The Per Protocol Set for Part B booster consisted of all Part B booster participants who had both pre-booster and post-booster immunogenicity assessments at (OL-Day 1 and OL-Day 29), did not have evidence of past or current SARS-CoV-2 infection at OL-Day 1 for Part B, where SARS-CoV-2 infection was defined as a positive RT-PCR test for SARS-CoV-2 and/or a positive serology test based on bAb specific to SARS-CoV-2 nucleocapsid (as measured by Roche Elecsys Anti-SARS-CoV-2 assay), and had no major protocol deviations that impacted immune response during the period corresponding to the immunogenicity analysis objective in Part B. The Per Protocol Immunogenicity Set for the booster in Part B served as the primary population for the analysis of immunogenicity data in Part B.

The geometric mean titers (GMT) of bAb or nAb titers with corresponding 95% CI were provided at each time point. The GM fold-rise (GMFR) of bAb or nAb titers and the corresponding 95% CI were also provided. The 95% CI was calculated based on the t-distribution of the log-transformed values or the difference in the log-transformed values for GMT and GMFR, respectively, then back transformed to the original scale for presentation. For calculation of GMTs and GMFRs, antibody values reported as below the LLOQ were replaced by 0.5 × LLOQ. Values that were greater than the upper limit of quantification (ULOQ) were converted to the ULOQ if actual values were not available. Missing results were not be imputed.

To assess the magnitudes of the differences in immune response 28 days after a single booster dose of 50 μg mRNA-1273 and the immune response 28 days after the completion of the primary series of mRNA-1273 100 μg in the phase 3 COVE study, an analysis of covariance (ANCOVA) model was used. The model included log-transformed antibody titers at D29 after booster in this phase 2 study, and D57 in COVE study as the dependent variable, treatment groups (50 μg mRNA-1273 booster in phase 2, 100 μg primary series in COVE) as explanatory variable, adjusting for age groups (< 65, ≥ 65; age group used in COVE Study). The geometric least squares mean (GLSM) and corresponding 2-sided 95% CI for the antibody titers for each treatment group were provided. The GLSM, and the corresponding 95% CI results in log-transformed scale estimated from the model were back-transformed to obtain these estimates in the original scale. GMR, estimated by the ratio of GLSM and the corresponding 2-sided 95% CI were provided to assess the treatment difference.

The primary immunogenicity objective in Part B was considered met if the noninferiority based on both GM titers and seroresponse rate at 28 days after the booster in Part B compared with 28 days after the second dose in the phase 3 COVE trial was demonstrated, at a 2-sided alpha of 0.05. The null hypotheses based on GM titers and seroresponse rate and the criterion of success included: Noninferiority based on the GMR (28 days after the booster in Part B versus 28 days after the second dose in the phase 3 COVE trial) was predefined with a noninferiority margin of 1.5 and a point estimate of GMR ≥ 1; and noninferiority based on difference in seroresponse rate (28 days after the booster in Part B minus 28 days after the second dose in the phase 3 COVE trial) was predefined with a noninferiority margin of 10%.

All analyses were conducted using SAS Version 9.4 or higher.

## Results

### Trial Population

The Phase 2 Part A trial consisted of a total of 600 participants. Of the 344 participants who received a booster dose in Part B, 173 received two doses of 50 µg of mRNA-1273 and 171 received two doses of 100 µg of mRNA-1273 6 to 8 months earlier in Part A (Figure 1).

The baseline demographic characteristics of the participants who received a booster injection were generally similar between the groups that received a primary series of 50 µg or 100 µg of mRNA-1273 and the group in the phase 3 COVE that received 2 doses of mRNA-1273 (Table 1). The majority of participants were White and not Hispanic or Latinx in the P201 and COVE trials, but there were higher percentages of Black and Hispanic or Latinx participants in the latter. The mean age of the participants in the groups that received the booster was 52.0 years in P201 Part B and 54.5 years for those who received 2 doses in the phase 3 COVE trial.

**Table 1.**
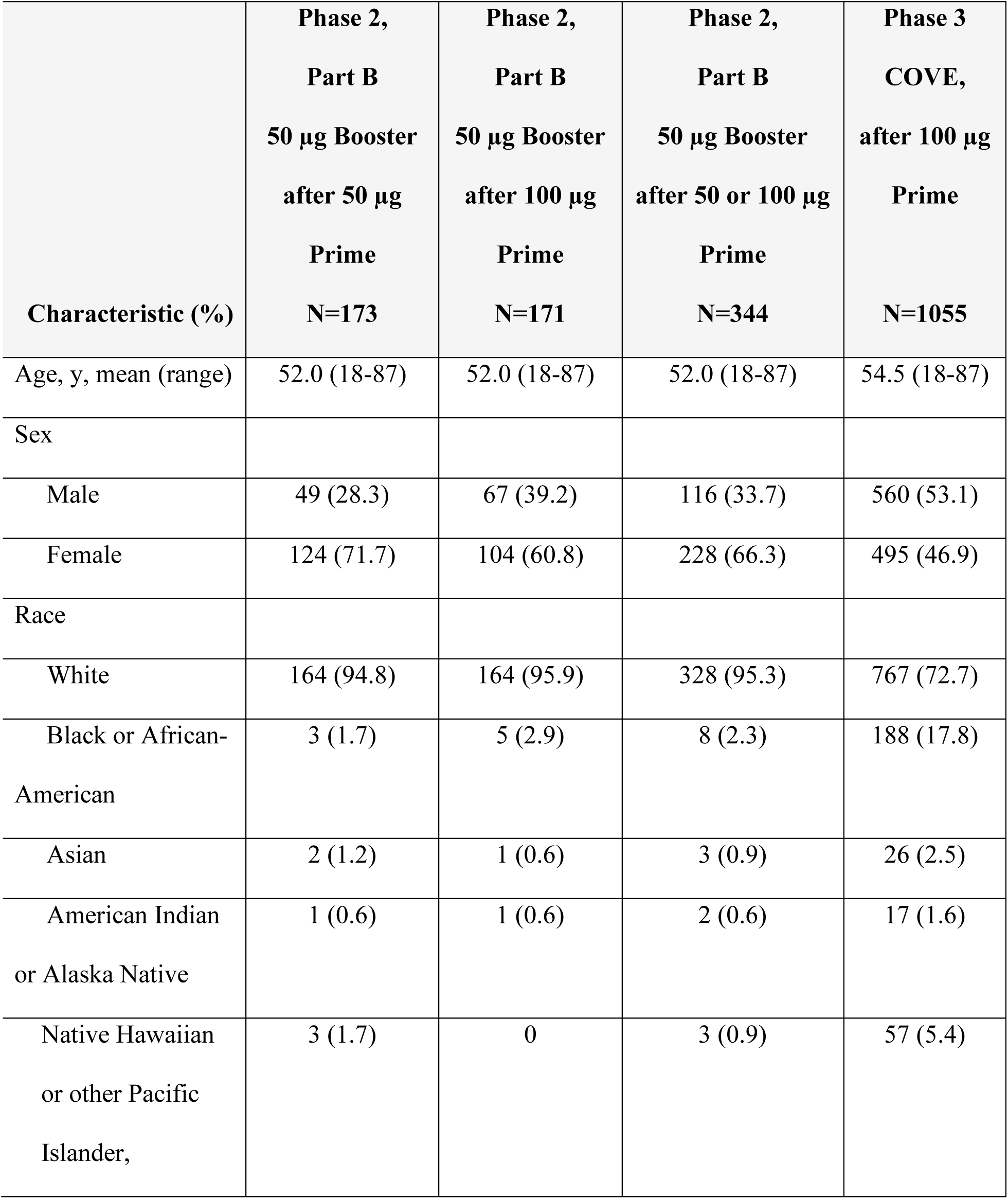
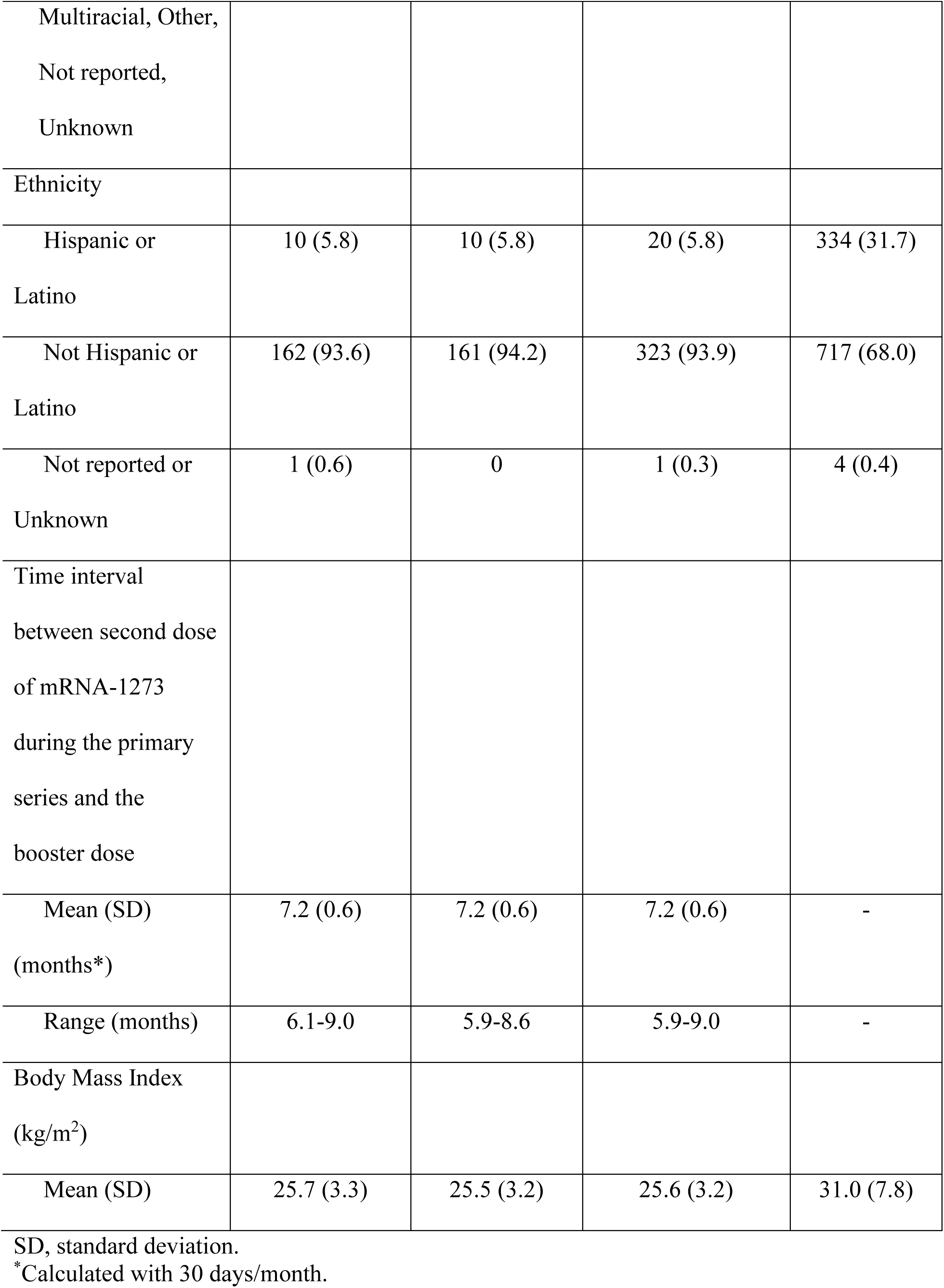
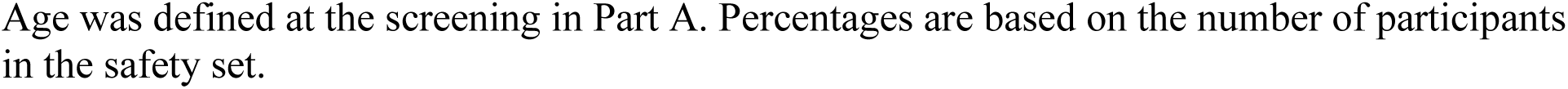
Demographics and Characteristics (Safety Set)

The time intervals (mean [standard deviation; range]) between the second dose of mRNA-1273 during the primary series and the booster injection was 7.2 [0.6; 6.1-9.0] months for the group that received 50 µg of mRNA-1273 and 7.2 [0.6; 5.9-8.6] months for the group that received 100 µg of mRNA-1273 during the primary series.

### Safety

The percentages of participants with any solicited local or systemic adverse reactions within seven days of the last injection were generally similar between the group that received a booster injection after a primary series of 50 µg or 100 µg of mRNA-1273 (Part B), the group in the phase 3 COVE trial that received two doses of 100 µg of mRNA-1273, and the group in this trial during the blinded phase (Part A) after they received two doses of mRNA-1273 (Figures 2A and 2B; Supplementary Table 1; Supplementary Table 2). The majority of the solicited local or systemic adverse reactions were mild (Grade 1) or moderate (Grade 2) (Figures 2A and 2B). The incidence of any Grade 3 solicited local or systemic adverse reaction after the booster injection were low (4.8%-12.9%) (Supplementary Table 1; Supplementary Table 2). There were no Grade 4 solicited local or systemic adverse events after the booster injection.

**Figure 2:**
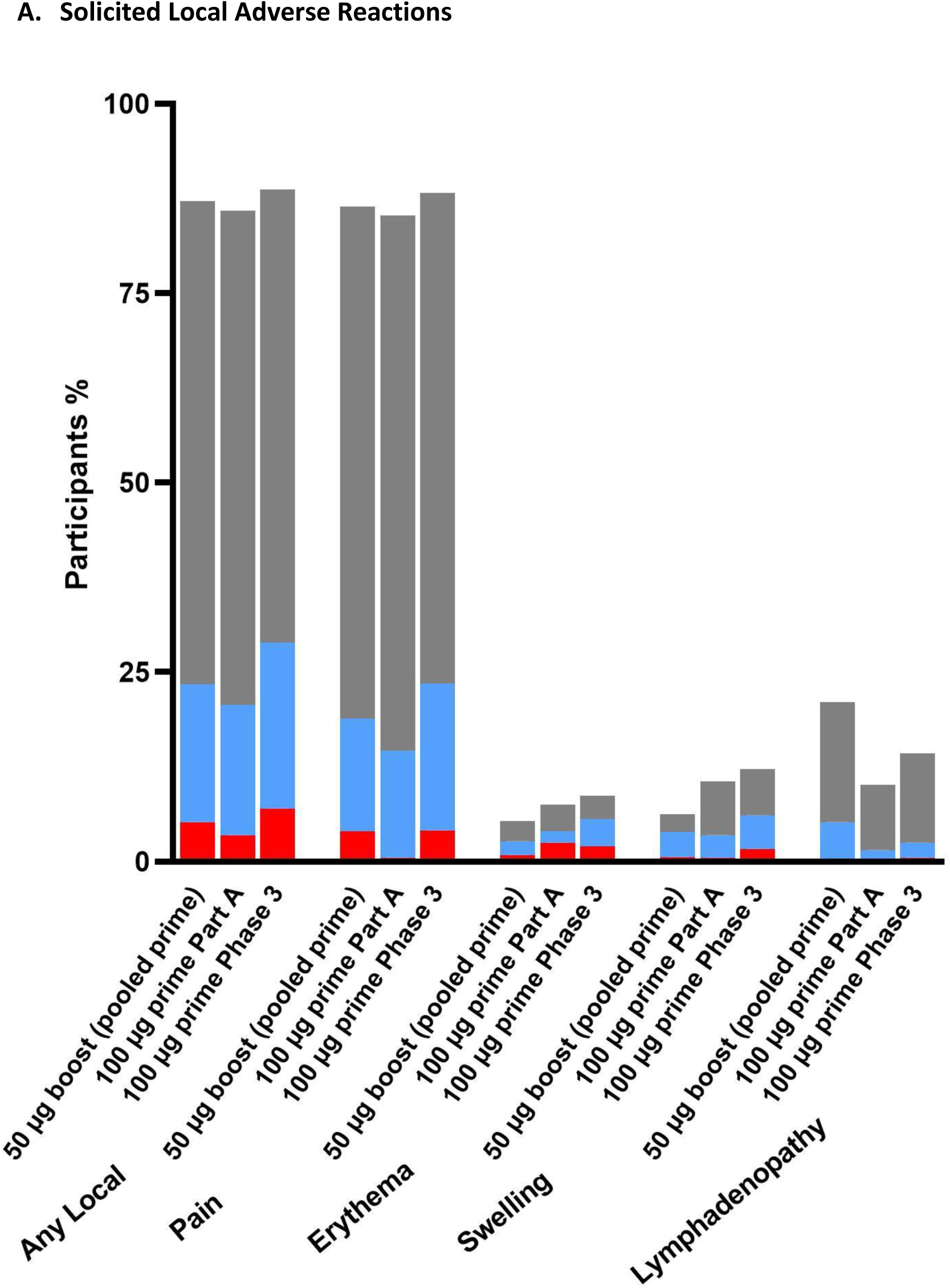
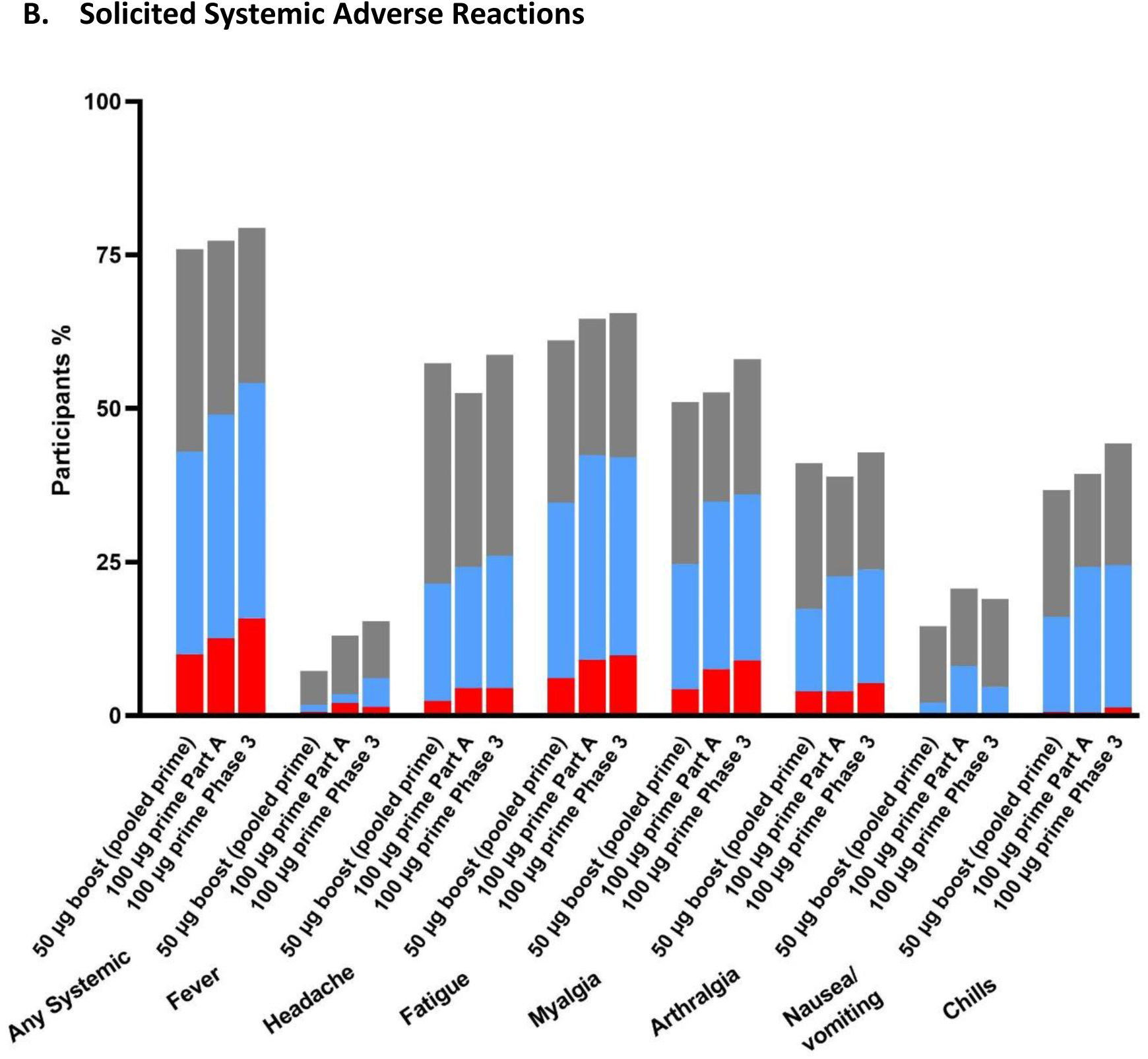
Solicited Adverse Reactions Within 7 Days After Booster Injection. Legend: The percentage of participants in the Solicited Safety Set who reported local (A) or systemic (B) solicited adverse reactions is shown for 330 participants who received a booster dose of mRNA-1273 (50 μg) after a primary series of two doses of 50 or 100 µg of mRNA-1273 in Part B, 198 participants who received a booster dose of mRNA-1273 (50 μg) after a primary series of two doses of 100 µg of mRNA-1273 in Part B, and 14691 participants who received two doses of 100 µg of mRNA-1273 in the phase 3 COVE trial. The percentages of participants who submitted any data for the adverse event within seven days after the booster injection or the second dose during the primary series are shown.

The most common local adverse reaction was injection-site pain in 86.3% of those in the pooled group that received the 50 µg and 100 µg prime and 88.3% of those in the phase 3 COVE trial (Supplementary Table 1). The most common Grade 3 local adverse reaction was injection-site pain in 4.0% of the participants in the pooled 50 and 100 µg prime group and 4.1% of those in the phase 3 COVE trial (Supplementary Table 1). Incidences of local solicited adverse reactions in participants who received a booster injection following a primary series of 50 or 100µg mRNA-1273 (Part B) were generally numerically similar to the participants’ local solicited adverse reactions after receiving a primary series of 50 or 100µg mRNA-1273 in this phase 2 trial (Part A), and participants in the phase 3 COVE trial who received only a primary series of 100 µg of mRNA-1273 (Supplementary Table 1; Figure 2A). However, lymphadenopathy was reported in 21.0% of participants who received a booster (pooled 50 and 100 µg prime) compared to 10.1% of participants in Part A who received two doses of 100 µg of mRNA-1273 and 14.2% of participants in the phase 3 COVE trial after receiving the second dose of mRNA-1273 (Supplementary Table 1).

The most common systemic adverse reactions after the booster dose of mRNA-1273 were fatigue, headache and myalgia (Figure 2B). Most solicited systemic adverse reactions after the booster injection were mild (Grade 1) or moderate (Grade 2) (Supplementary Table 2; Figure 2B). The most common Grade 3 systemic adverse reaction after the booster dose was fatigue in 6.1% of the participants (Figure 2B; Supplementary Table 2). The incidences of systemic solicited adverse reactions were numerically similar in the pooled group that received a booster injection after a primary series of 50 µg or 100 µg of mRNA-1273, the group in phase 2 Part A that received two doses of 100 µg of mRNA-1273 or the group in phase 3 COVE trial after receiving the second dose of 100 µg of mRNA-1273 (Figure 2B; Supplementary Table 2).

The percentages of participants with any solicited local or systemic adverse reactions were generally similar between the groups that received a booster injection and were ≥18 to <55 years of age compared to those who were ≥55 years of age (Supplementary Table 3). Thirteen participants (3.8%) reported treatment-emergent adverse events related to study vaccination up to 28 days after the booster injection, but none led to study discontinuation (Supplementary Table 7). There were no serious adverse events up to 28 days after the booster injection.

### Immunogenicity

The GMTs [95% confidence interval] in the D614G pseudovirus neutralizing antibody assay at Day 57 (28 days after the second injection of mRNA-1273 during the primary series) were 896.5 (804.4, 1000.4) in the pooled group who received 50 or 100 µg of mRNA-1273, 629.2 (549.3, 720.8) in the 50 µg group and 1268.0 (1087.9, 1477.8) in the 100 µg group (Figure 3; Supplementary Table 4). Before the administration of the booster (OL-Day 1; Pre-boost), the levels of neutralizing antibodies were 104.7 (88.3, 124.1) and 150.2 (125.7, 179.5) in the 50 µg group and 100 µg group, respectively (Figure 3; Supplementary Table 4). At OL-Day 29 (28 days after the 50 µg booster), the levels of neutralizing antibody titers were 1834.3 (1600.2, 2102.6) in the 50 µg group and 1951.7 (1729.6, 2202.4) in the 100 µg group (Figure 3, Supplementary Table 4). The GMTs (95% CIs) of neutralizing antibody at 28 days after the booster [1834.3 (1600.2, 2102.6) in the 50 µg group and 1951.7 (1729.6, 2202.4) in the 100 µg group] were higher than the levels of antibody at 28 days after the second dose of mRNA-1273 during the primary series [629.2 (549.3, 720.8) in the 50 µg group and 1268.0 (1087.9, 1477.8) in the 100 µg group]. The GM fold rises at 28 days after the booster compared to 28 days after the second dose in the primary series were 2.1 (1.9, 2.3) for the pooled group of 50 and 100 µg, 2.9 (2.6, 3.4) for the 50 µg group and 1.5 (1.3, 1.8) for the 100 µg group (Supplementary Table 4). The difference in the GM fold rises between the 50 and 100 µg groups is most likely due to differences in the GMTs in these 2 groups at one month after the second injection of mRNA-1273 (Figure 3; Supplementary Table 4).

**Figure 3:**
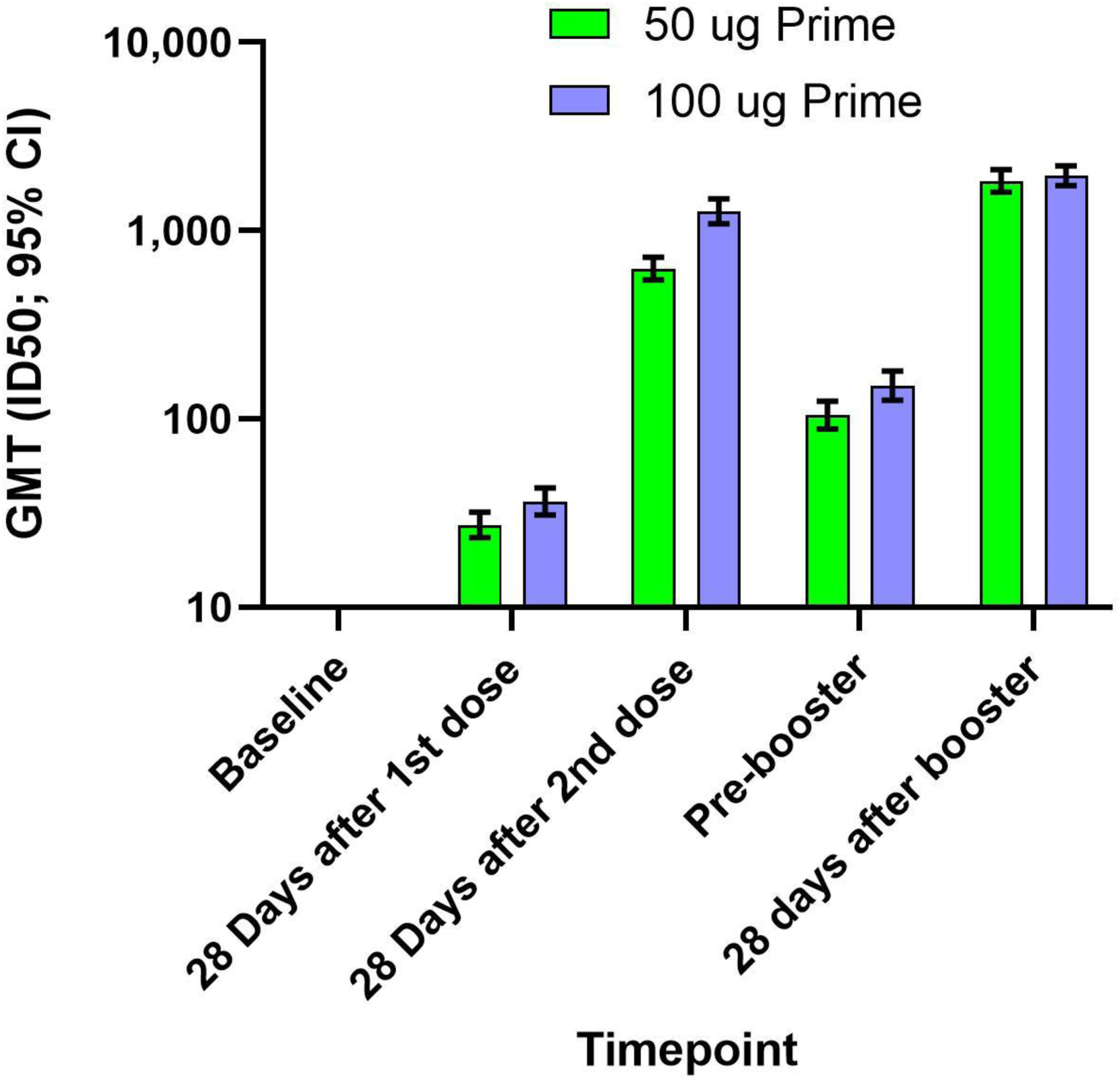
Neutralizing Antibody Titers (Pseudovirus ID50; D614G) After the Primary Series and After a Booster Injection of 50 µg of mRNA-1273 (Per-protocol Set) The geometric mean titers (GMTs) and 95% confidence intervals (95% CIs) against the D614G virus for serum samples collected in Part A at baseline, 28 days after the first dose of mRNA-1273, 28 days after the second dose of mRNA-1273, and in Part B before the booster injection of 50 µg of mRNA-1273 (Pre-booster) and 28 days after the booster injection. Antibody values in the pseudovirus assay reported as below the lower limit of quantification (LLOQ; 18.5) were replaced by 0.5 x LLOQ. Values that were greater than the upper limit of quantification (ULOQ; 45118) were changed to the ULOQ if actual values were not available. 95% Confidence Intervals were calculated based on the t-distribution of the log-transformed values or the difference in the log-transformed values for GMT, then back transformed to the original scale.

The neutralizing antibody levels at 28 days after the booster dose of mRNA-1273 in participants in Part B of this study were also higher than those at 28 days after the second dose of mRNA-1273 in participants in the pivotal phase 3 COVE trial (Table 2). In a comparison of the pseudovirus neutralizing antibody titers against D614G at 28 days after the booster dose with those in the phase 3 COVE trial at 28 days after the second dose (in which vaccine efficacy was demonstrated^5^), the geometric mean ratio (GMR) was 1.7 (95% CI: 1.5, 1.9) (Table 2). This GMR of 1.7 was above the prespecified threshold of 1.0 and the lower bound of the 95% CI was greater than 0.67 (corresponding to a noninferiority margin of 1.5). Therefore, the prespecified criteria for noninferiority were met for both the GMT ratio and the seroresponse difference. In a comparison of the pseudovirus neutralizing antibody titers against the Delta variant at 28 days after the booster dose with those in the phase 3 COVE trial at 28 days after the second dose^30^, the geometric mean ratio (GMR) was 2.1 (95% CI: 1.8, 2.4) (Table 2). The higher antibody levels at 28 days after the booster dose compared to 28 days after the second dose in the phase 3 COVE study were observed in three assays: pseudovirus neutralizing antibodies, anti-spike IgG antibody by ELISA (VAC65) and anti-spike IgG antibody by MSD MULTIPLEX (Supplementary Table 5).

**Table 2:** Pseudovirus Neutralizing Antibody Titers (ID50; Against D614G or Delta) of mRNA-1273 Post-Booster Compared with the Phase 3 COVE Primary Series Titers (Per Protocol Immunogenicity Set)

| **Pseudovirus Neutralization Assay** | **D614G** |  | **Delta (B.1.617.2)** |  |
| --- | --- | --- | --- | --- |
|  | **Phase 2, Part B,**<br>**50 µg mRNA-1273 Booster** | **Phase 3 COVE,**<br>**100 µg mRNA-1273**<br>**Primary Series** | **Phase 2, Part B,**<br>**50 µg mRNA-1273 Booster** | **Phase 3 COVE,**<br>**100 µg mRNA-1273**<br>**Primary Series** |
| **28 Days After Booster (OL-D29; Phase 2 Part B) or 28 Days After Second Dose in Phase 3 COVE** |  |  |  |  |
| **n** | 295 | 1053 | 295 | 580 |
| **GLSM** | 1767.9 | 1032.7 | 743.9 | 354.0 |
| **95% CI** | 1586.4, 1970.2 | 974.2, 1094.7 | 663.7, 833.7 | 325.0, 385.5 |
| **GMR (Phase 2 Part B vs. Phase 3 COVE; model-based)** | 1.7 |  | 2.1 |  |
| **95% CI** | 1.5, 1.9 |  | 1.8, 2.4 |  |

Abbreviations: ANCOVA = analysis of covariance; ID50 = 50% inhibitory dilution; GLSM = geometric least squares mean; CI = confidence interval; GMR = geometric mean ratio; LLOQ = lower limit of quantification; ULOQ = upper limit of quantification.
n= number of subjects with no missing data at the corresponding timepoint. Antibody values reported as below the LLOQ were replaced by $0.5 \times \text{LLOQ}$ . Values greater than the ULOQ were replaced by the ULOQ if actual values are not available.
The log-transformed antibody levels were analyzed using an analysis of covariance (ANCOVA) model with the group variable (P201 Part B and P301) as fixed effect. The resulted LS means, difference of LS means, and 95% CI were back transformed to the original scale for presentation.
Source: Table 2.1 and 2.5

Seroresponse rates (assay-specific definition) [95% CI] were 93.5% [90.1, 96.1] and 98.9% [98.0, 99.4] in the pooled Phase 2 Part B booster group (compared to OL-D1 pre-boost) and after the primary series in the phase 3 COVE trial, respectively (Table 3). The seroresponse rates (4-fold definition) [95% CI] were 90.1% [86.1, 93.3] and 98.4% [97.4, 99.1] in the booster group and after the primary series in the phase 3 COVE trial, respectively (Table 3). The seroresponse rates (4-fold rise from baseline) [95% CI] were 100% [98.7, 100.0] and 98.3% [96.0, 99.4] in the pooled Phase 2 Part B booster group and in the pooled Phase 2 Part A priming group (50 µg or 100 µg), respectively (Table 3). The seroresponse rates in the pseudovirus neutralizing antibody assay were noninferior in the phase 2 Part B group after the booster injection compared to the phase 3 COVE trial after the primary series (Table 3), based on the assay-specific seroresponse definition. Given that the lower limit of the 95% confidence interval for the group difference in seroresponse rates (4-fold rise from baseline) was greater than 0, the observed seroresponse rates are statistically significantly higher after the booster dose than after the second dose in Part A of the phase 2 study.

**Table 3:**
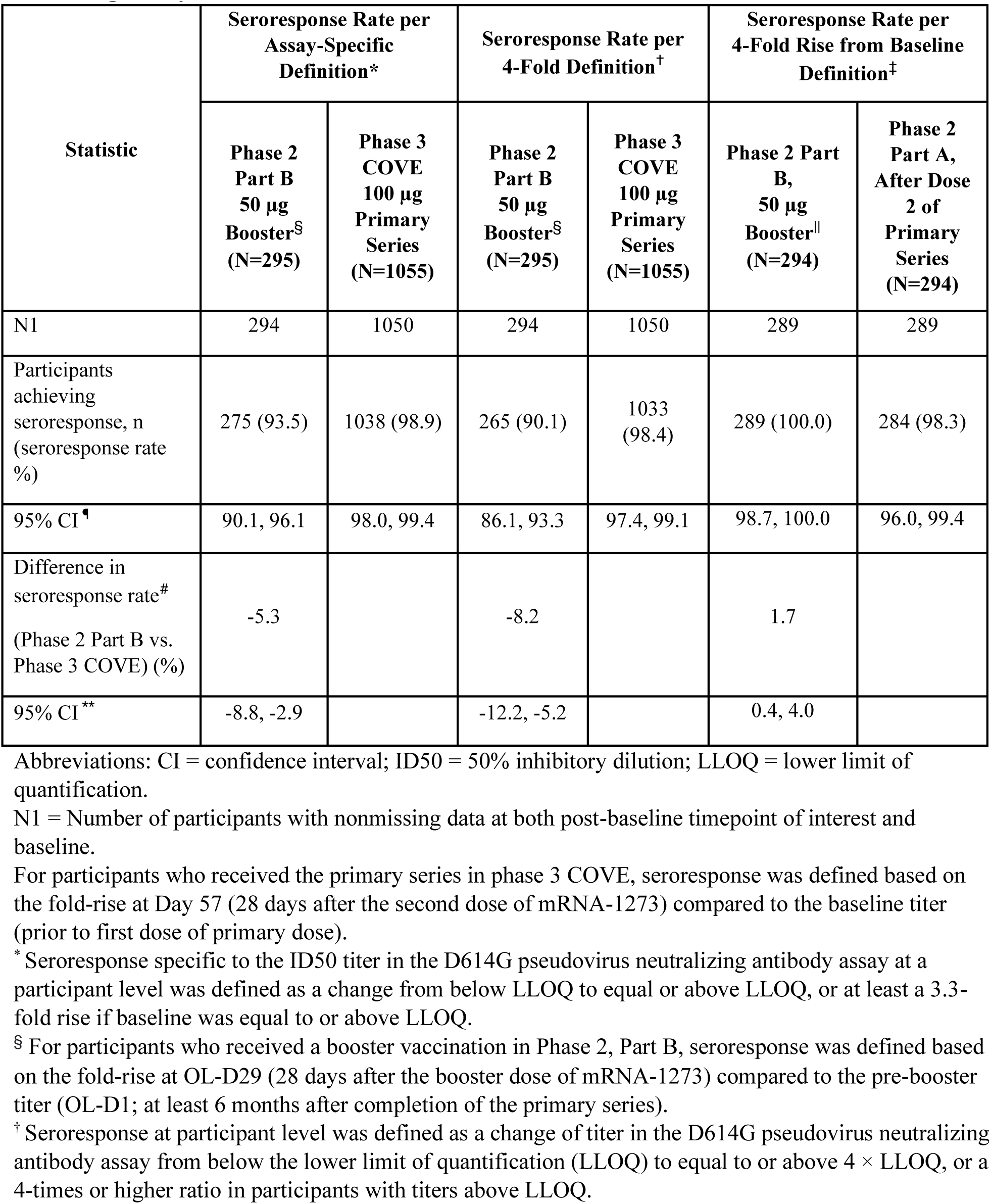
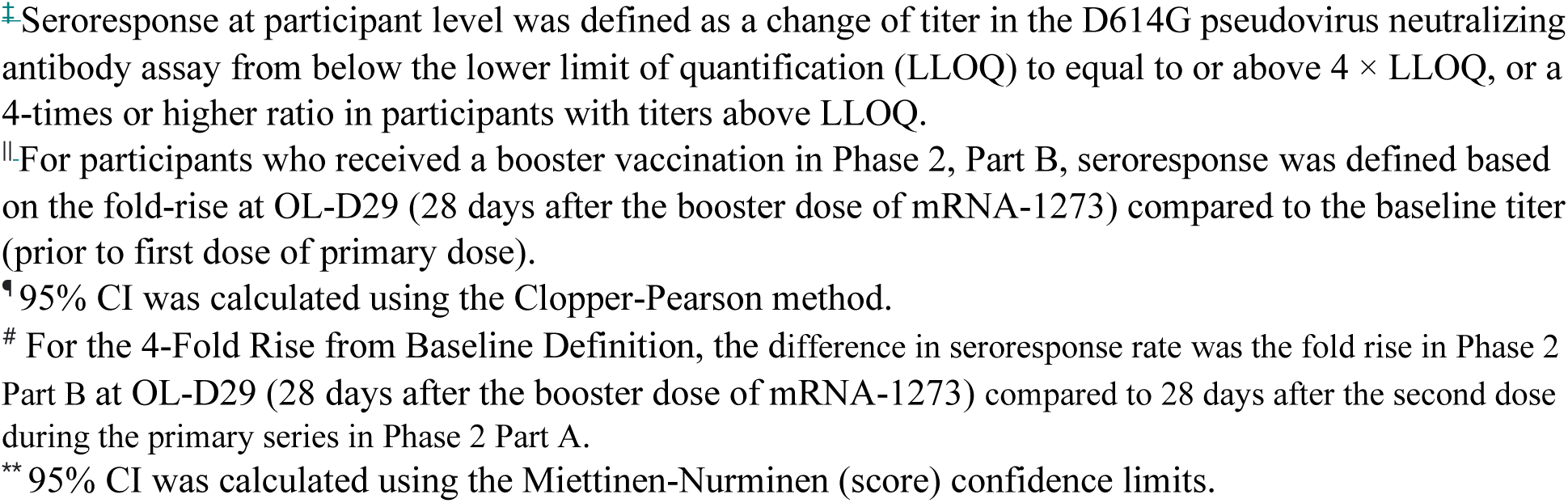
Seroresponse Rates by Pseudovirus Neutralizing Antibody (ID50; D614G) Assay: Phase 2 after Booster Compared with the Phase 3 COVE Primary Series (Per Protocol Immunogenicity Set)

In the pooled Part B group (previously primed with 2 doses of either 50 or 100 ug mRNA-1273, n=293), the pre-booster nAb GMT versus the Delta variant was 42.3 (95% CI, 37.2, 48.0; n=293) and the GMT at 28 days post-booster was 803.5 (95% CI, 731.4, 882.7; n=295) (Supplementary Table 8). Over 90% of booster recipients in the overall group (92.2%; 95% CI, 88.5-95.0%; n=293) met the definition of a seroresponse to the Delta variant using a four-fold increase from pre-booster baseline. Administration of the mRNA-1273 booster (50 ug) induced a 19-fold-rise in neutralizing titers against the Delta variant compared to pre-booster levels (GMFR=19.0; 95% CI, 16.7, 21.5; pooled group, n=295; Supplementary Table 8). Titers in the pooled group 28 days after the booster dose were 2.4-fold lower against the Delta variant (803.5; Supplementary Table 8) than against the D614G virus (1892.7; Supplementary Table 4), which is similar to the 2.9-fold difference (GMTs=354 against Delta, and 1032.7 against D614G; Table 2) seen 28 days after the primary series of two doses of 100 µg mRNA-1273 in the phase 3 COVE study.

## Discussion

In this study, the administration of a booster dose of 50 µg mRNA-1273 to participants approximately 6 to 8 months after a primary series of two doses of 50 or 100 µg of mRNA-1273 resulted in a safety profile that was comparable to that in the group in the phase 3 COVE trial after receiving the second dose of mRNA-1273 in the primary series. After receiving the booster injection of mRNA-1273, the most common local adverse reaction was pain, and the most common systemic adverse reactions were fatigue, headache and myalgia. The incidence and severity of local and systemic adverse reactions was similar to that observed in Part A of this study and the phase 3 COVE trial after the second injection of mRNA-1273.^5, 11^ Both co-primary immunogenicity objectives were met for the pooled group that received a booster (Part B) compared to the group in the phase 3 COVE trial that received two doses of mRNA-1273. Neutralizing antibody titers (pseudovirus neutralizing antibody) against the D614G strain of virus at 28 days after the booster injection were higher than the levels at 28 days after the second dose of mRNA-1273 during the primary series in the pivotal phase 3 efficacy trial (COVE study). This higher level of antibody after the booster injection compared to the second injection is indicative of a robust immune memory response likely due to stimulation of memory B cells.^30–33^ It is also important to note that the results here using a validated pseudovirus neutralization assay showed that the GMTs after the second dose of mRNA-1273 were higher at one month after the second dose in the 100 µg group (1268.0; 95% CI=1087.9, 1477.8) compared to the 50 µg group (629.2; 549.3, 720.8) (Figure 3; Supplementary Table 4). Previous results using a qualified live virus neutralizing antibody assay showed similar GMTs at one month after the second dose of 100 µg compared to 50 µg (∼1640).^28^ These different results between the 100 µg and 50 µg groups are most likely due to differences in the dynamic ranges of the neutralizing antibody assays.

SARS-CoV-2 vaccines appear to have retained their effectiveness in preventing hospitalization and severe disease but the efficacy to prevent asymptomatic infection or mild symptomatic disease may have decreased since the emergence of the Delta variant.^24, 27, 34–36^ The potential reasons for the lower vaccine effectiveness include both waning immunity and the emergence of viral variants which may be more transmissible than the original Wuhan-1 strain. In preliminary results at 6 months after two doses of mRNA-1273, neutralizing antibody levels against the Delta variant decreased compared to 28 days after the second dose of mRNA-1273 in the primary series, and approximately one-half of participants had no detectable neutralizing antibodies against the Delta variant in a pseudovirus research grade assay.^23^ At 2 weeks after a booster dose of 50 µg of mRNA-1273, neutralizing antibody levels to the Delta variant increased to a similar level as neutralizing antibody titers against wild-type D614G at one month after the second dose of mRNA-1273 in the primary series.^23^

The GMFR (Day 29 post-booster compared to pre-booster) achieved by mRNA-1273 booster, measured by the Delta variant pseudovirus assay (18.97; 95% CI, 16.72, 21.53), points to the ability of the mRNA-1273 vaccine booster to enhance a breadth of neutralizing antibody responses, including against the highly transmissible Delta variant. Just as booster mRNA-1273 stimulated nAb levels against the original strain (GMTs 1892.7 at OL-D29 compared to 125.7 at OL-D1 (Supplementary Table 4), a booster injection of mRNA-1273 was able to broaden and increase nAb levels against the Delta variant, highlighting the critical benefits of mRNA-1273 booster dose.

There are several limitations to the results of this study. This study was designed to assess the safety and immune response of a third dose of mRNA-1273 administered at least 6 months after the initial priming series. While the optimal timing of a third dose has not been established, the data from this trial provides important information to address the need for third booster dose in case of waning vaccine effectiveness. Although neutralizing antibody responses have been correlated to reduction of risk for breakthrough COVID-19 disease,^37^ a threshold of protection has not been defined for the Wuhan-1 or Delta variants. Also, this study did not examine variant-specific booster vaccines or immune responses to variants of concern other than for Delta. Finally, this study showed an immune memory antibody response to the spike protein of SARS-CoV-2 but did not examine T cell memory or quantify B memory cells.

While the data supporting the timing of when booster doses of mRNA vaccine against SARS-CoV-2 should be administered is still evolving, the results from this study provide evidence that a third dose of mRNA-1273 administered at least 6 months after the primary series is safe and effective in inducing a booster response, as indicated by the statistically significantly higher antibody titers observed after the 50 µg booster dose as compared to after the second priming dose of 100 µg of mRNA-1273. A booster dose of mRNA-1273 has the potential for establishing durable vaccine efficacy and restoring neutralizing antibody capability. The increased antibody responses to the Delta variant suggest that a third dose of mRNA-1273 may provide improved protection against this variant of concern.

## Data Availability

The data supporting the findings of this study are available in this article and in its Supplementary Information.

## Acknowledgements

We thanks the participants in this trial and the phase 2 Study Investigators; the Immune Assay Team at Duke University Medical Center: Rebecca Beerman, Kendall Bradley, Jiayu Chen, Xiaoju Daniell, Elizabeth Domin, Amanda Eaton, Kelsey Engle, Wenhong Feng, Juanfei Gao, Hongmei Gao, Kelli Greene, Sarah Hiles, Marianne Jessup-Cumming, Marcella Sarzotti-Kelsoe, Kristy Long, Kellen Lund, Kaia Lyons, Charlene McDanal, Francesca Suman, Haili Tang, Jin Tong, Olivia Widman; the mRNA-1273 Product Coordination Team from the Biomedical Advanced Research and Development Authority (BARDA) (Robert Bruno, Richard Gorman, Holli Hamilton, Gary Horwith, Chuong Huynh, Nutan Mytle, Corrina Pavetto, Xiaomi Tong, and John Treanor); the team at PPD, Hamilton, New Jersey, for their support of this study; and Frank J. Dutko, Ph.D. and Joanne E. Tomassini, Ph.D., Moderna consultants, for contributions to writing the manuscript.

## Supplementary Information

**Supplementary Figure 1:**
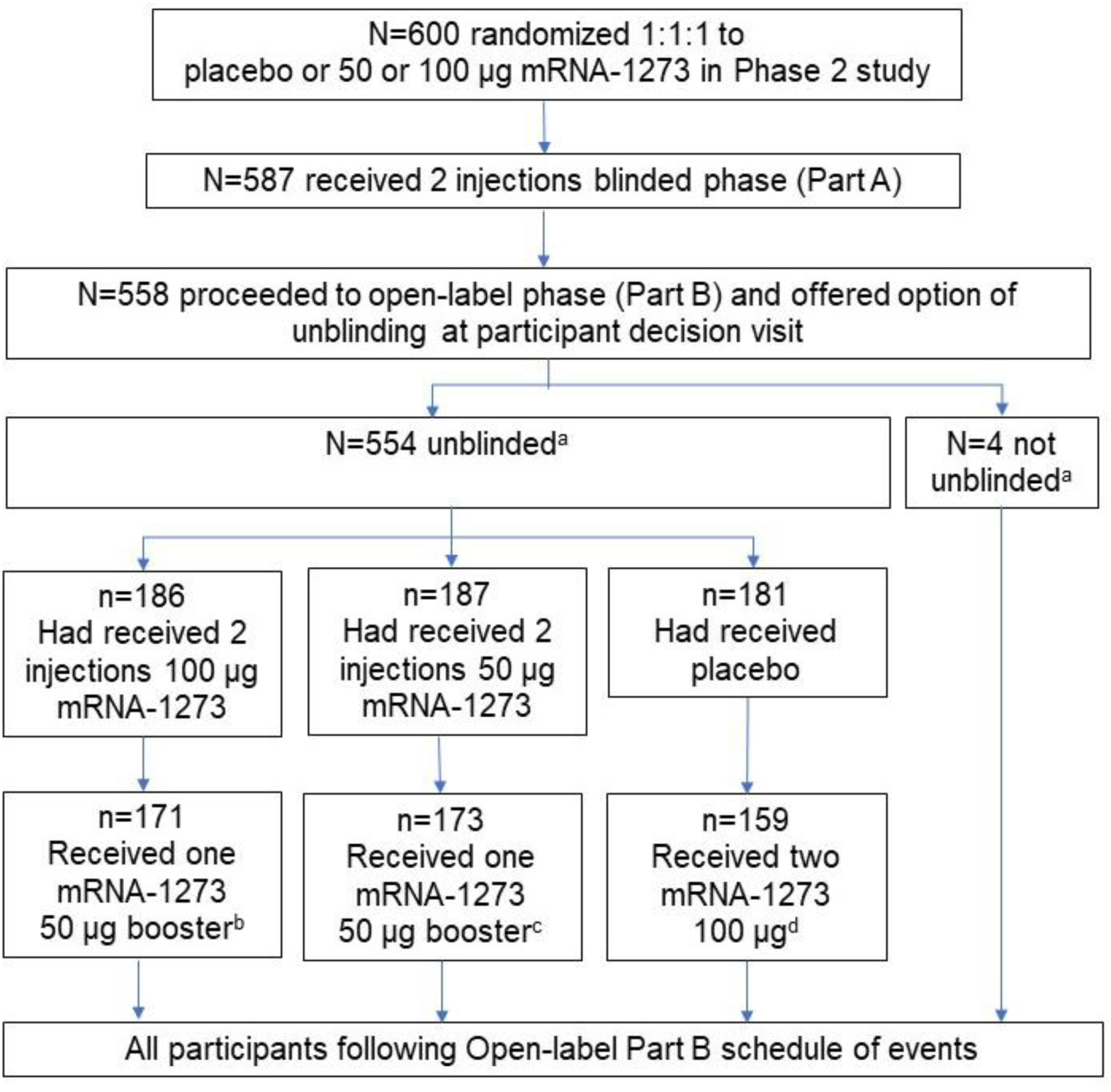
Participant Flow Diagram. In Part B, participants who had received 2 injections of 50 μg or 100 μg mRNA-1273 or placebo completed the blinded phase (Part A) and went on to receive a single open-label booster dose of 50 μg mRNA-1273 or two doses of 100 µg of mRNA-1273. ^a^Unblinded or not unblinded to assigned treatment in Part A blinded phase. ^b^15 participants declined to receive a booster of 50 μg mRNA-1273. ^c^14 participants declined to receive a booster of 50 μg mRNA-1273. ^d^22 participants declined to receive mRNA-1273 in Part B.

**Supplementary Figure 2:**
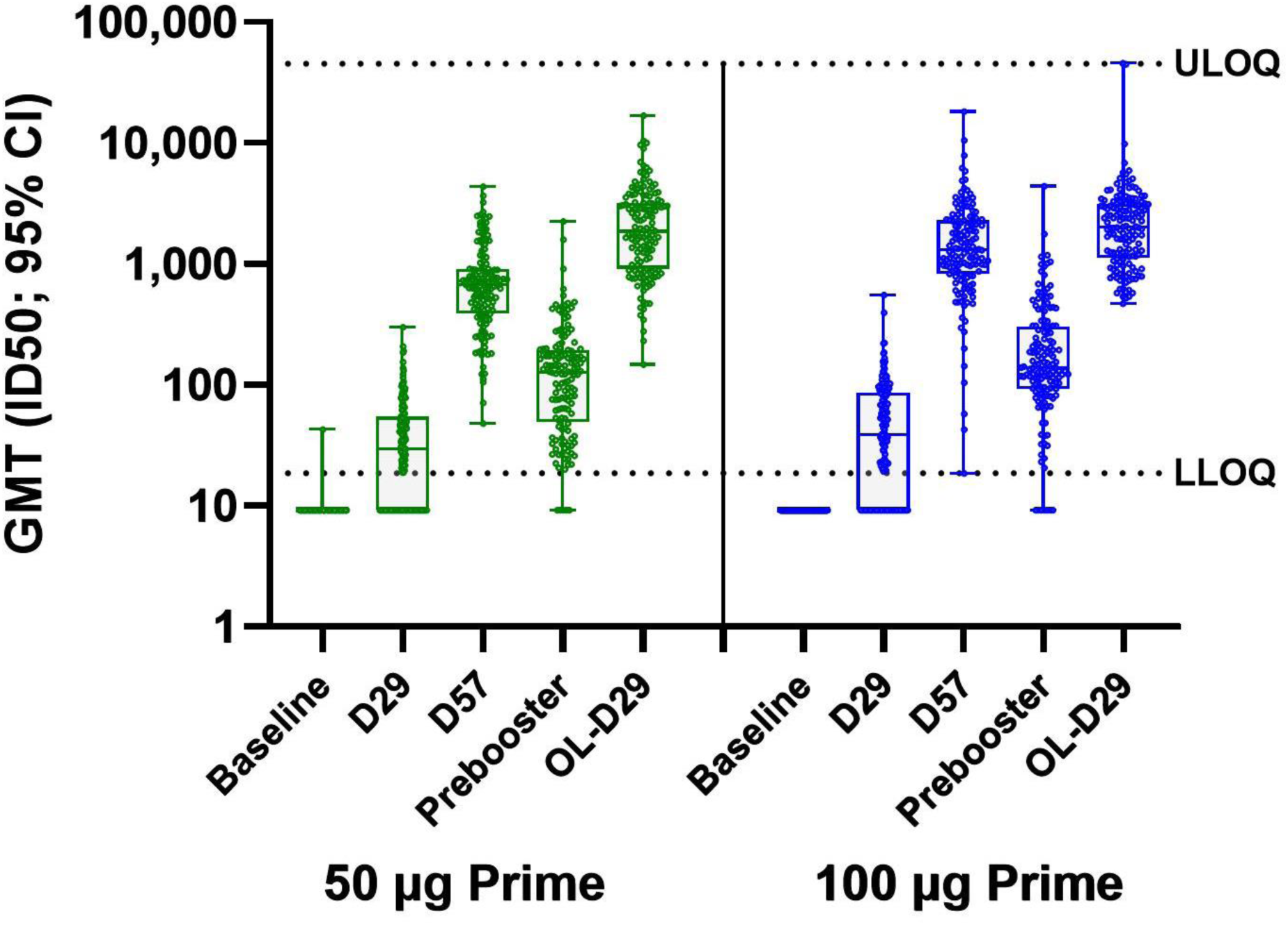
Neutralizing Antibody Titers (Pseudovirus ID50; D614G) After the Primary Series and After a Booster Injection of 50 µg of mRNA-1273 (Per-protocol Set) Boxes indicate interquartile ranges (IQR) from Q1 to Q3 of the D614G neutralizing antibody values in the Per-protocol Immunogenicity Subset at various time points during the primary series (Baseline, D29 (28 days after the first dose of mRNA-1273), D57 (28 days after the second dose of mRNA-1273, Pre-booster (OL-Day 1), and OL-D29 (28 days after the booster dose of 50 µg of mRNA-1273). The medians are shown by the horizontal lines within the boxes. The error bars (whiskers) show the minimum to the maximum antibody values. Individual data points are shown by the circles. Antibody values reported as below the lower limit of quantification (LLOQ; 18.5) were replaced by 0.5 x LLOQ. Values that were greater than the upper limit of quantification (ULOQ; 45118) were changed to the ULOQ if actual values were not available.

---

**Supplementary Table 1:**
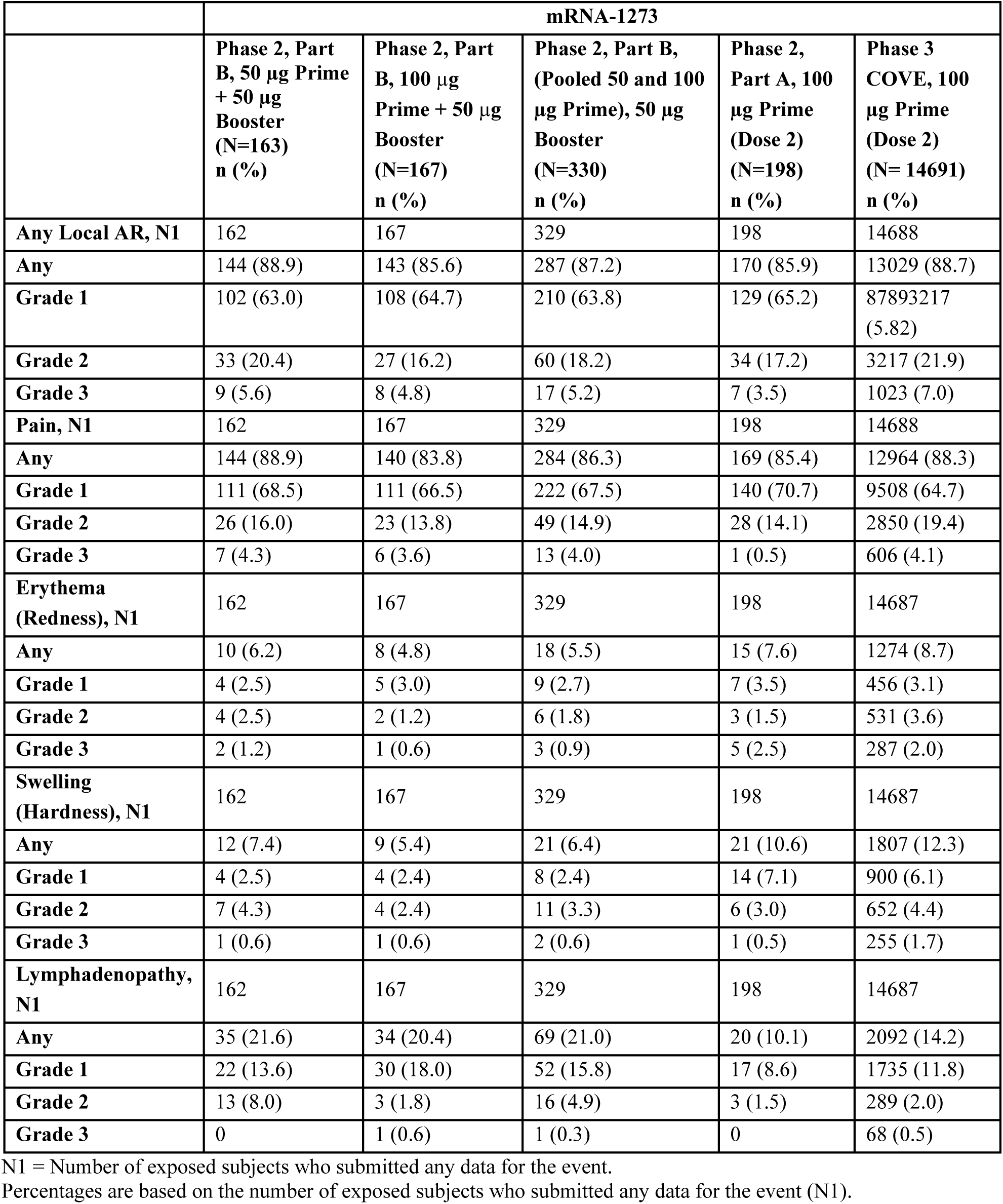
Solicited Local Adverse Reactions Reported Within 7 Days After 50 µg Booster in Phase 2 versus After the 2^nd^ Injection in the Primary Series of Phase 2 Part A or Phase 3 COVE (Solicited Safety Set)

|  | mRNA-1273 |  |  |  |  |
| --- | --- | --- | --- | --- | --- |
|  | Phase 2, Part B, 50 µg Prime + 50 µg Booster (N=163)<br>n (%) | Phase 2, Part B, 100 µg Prime + 50 µg Booster (N=167)<br>n (%) | Phase 2, Part B, (Pooled 50 and 100 µg Prime), 50 µg Booster (N=330)<br>n (%) | Phase 2, Part A, 100 µg Prime (Dose 2) (N=198)<br>n (%) | Phase 3 COVE, 100 µg Prime (Dose 2) (N= 14691)<br>n (%) |
| **Any Local AR, N1** | 162 | 167 | 329 | 198 | 14688 |
| **Any** | 144 (88.9) | 143 (85.6) | 287 (87.2) | 170 (85.9) | 13029 (88.7) |
| **Grade 1** | 102 (63.0) | 108 (64.7) | 210 (63.8) | 129 (65.2) | 87893217 (5.82) |
| **Grade 2** | 33 (20.4) | 27 (16.2) | 60 (18.2) | 34 (17.2) | 3217 (21.9) |
| **Grade 3** | 9 (5.6) | 8 (4.8) | 17 (5.2) | 7 (3.5) | 1023 (7.0) |
| **Pain, N1** | 162 | 167 | 329 | 198 | 14688 |
| **Any** | 144 (88.9) | 140 (83.8) | 284 (86.3) | 169 (85.4) | 12964 (88.3) |
| **Grade 1** | 111 (68.5) | 111 (66.5) | 222 (67.5) | 140 (70.7) | 9508 (64.7) |
| **Grade 2** | 26 (16.0) | 23 (13.8) | 49 (14.9) | 28 (14.1) | 2850 (19.4) |
| **Grade 3** | 7 (4.3) | 6 (3.6) | 13 (4.0) | 1 (0.5) | 606 (4.1) |
| **Erythema (Redness), N1** | 162 | 167 | 329 | 198 | 14687 |
| **Any** | 10 (6.2) | 8 (4.8) | 18 (5.5) | 15 (7.6) | 1274 (8.7) |
| **Grade 1** | 4 (2.5) | 5 (3.0) | 9 (2.7) | 7 (3.5) | 456 (3.1) |
| **Grade 2** | 4 (2.5) | 2 (1.2) | 6 (1.8) | 3 (1.5) | 531 (3.6) |
| **Grade 3** | 2 (1.2) | 1 (0.6) | 3 (0.9) | 5 (2.5) | 287 (2.0) |
| **Swelling (Hardness), N1** | 162 | 167 | 329 | 198 | 14687 |
| **Any** | 12 (7.4) | 9 (5.4) | 21 (6.4) | 21 (10.6) | 1807 (12.3) |
| **Grade 1** | 4 (2.5) | 4 (2.4) | 8 (2.4) | 14 (7.1) | 900 (6.1) |
| **Grade 2** | 7 (4.3) | 4 (2.4) | 11 (3.3) | 6 (3.0) | 652 (4.4) |
| **Grade 3** | 1 (0.6) | 1 (0.6) | 2 (0.6) | 1 (0.5) | 255 (1.7) |
| **Lymphadenopathy, N1** | 162 | 167 | 329 | 198 | 14687 |
| **Any** | 35 (21.6) | 34 (20.4) | 69 (21.0) | 20 (10.1) | 2092 (14.2) |
| **Grade 1** | 22 (13.6) | 30 (18.0) | 52 (15.8) | 17 (8.6) | 1735 (11.8) |
| **Grade 2** | 13 (8.0) | 3 (1.8) | 16 (4.9) | 3 (1.5) | 289 (2.0) |
| **Grade 3** | 0 | 1 (0.6) | 1 (0.3) | 0 | 68 (0.5) |

N1 = Number of exposed subjects who submitted any data for the event.
Percentages are based on the number of exposed subjects who submitted any data for the event (N1).

**Supplementary Table 2:** Solicited Systemic Adverse Reactions Reported Within 7 Days After 50 µg Booster in Phase 2 versus After the 2^nd^ Injection in the Primary Series of Phase 2 Part A or Phase 3 COVE (Solicited Safety Set)

|  | mRNA-1273 |  |  |  |  |
| --- | --- | --- | --- | --- | --- |
|  | **Phase 2, Part B, 50 µg Prime + 50 µg Booster (N=163)**<br>**n (%)** | **Phase 2, Part B, 100 µg Prime + 50 µg Booster N=167**<br>**n (%)** | **Phase 2, Part B, 50 µg Booster (Pooled 50 and 100 µg Prime) (N=330)**<br>**n (%)** | **Phase 2, Part A, 100 µg Prime (Dose 2) N=198**<br>**n (%)** | **Phase 3 COVE, 100 µg Prime (Dose 2) N=14691**<br>**n (%)** |
| **Any Systemic AR, N1** | 163 | 167 | 330 | 198 | 14690 |
| **Any** | 127 (77.9) | 126 (75.4) | 253 (76.7) | 153 (77.3) | 11678 (79.5) |
| **Grade 1** | 49 (30.1) | 60 (35.9) | 109 (33.0) | 56 (28.3) | 3717 (25.3) |
| **Grade 2** | 56 (34.4) | 53 (31.7) | 109 (33.0) | 72 (36.4) | 5611 (38.2) |
| **Grade 3** | 21 (12.9) | 12 (7.2) | 33 (10.0) | 25 (12.6) | 2336 (15.9) |
| **Grade 4** | 0 | 0 | 0 | 0 | 14 (<0.1) |
| **Fever, N1** | 162 | 166 | 328 | 198 | 14682 |
| **Any** | 13 (8.0) | 11 (6.6) | 24 (7.3) | 38 (13.1) | 2276 (15.5) |
| **Grade 1** | 12 (7.4) | 6 (3.6) | 18 (5.5) | 19 (9.6) | 1363 (9.3) |
| **Grade 2** | 1 (0.6) | 3 (1.8) | 4 (1.2) | 3 (1.5) | 697 (4.7) |
| **Grade 3** | 0 | 2 (1.2) | 2 (0.6) | 4 (2.0) | 203 (1.4) |
| **Grade 4** | 0 | 0 | 0 | 0 | 13 (<0.1) |
| **Headache, N1** | 162 | 167 | 329 | 198 | 14687 |
| **Any** | 97 (59.9) | 92 (55.1) | 189 (57.4) | 104 (52.5) | 8637 (58.8) |
| **Grade 1** | 57 (35.2) | 61 (36.5) | 118 (35.9) | 56 (28.3) | 4815 (32.8) |
| **Grade 2** | 34 (21.0) | 29 (17.4) | 63 (19.1) | 39 (19.7) | 3156 (21.5) |
| **Grade 3** | 6 (3.7) | 2 (1.2) | 8 (2.4) | 9 (4.5) | 666 (4.5) |
| **Fatigue, N1** | 162 | 167 | 329 | 198 | 14687 |
| **Any** | 103 (63.6) | 98 (58.7) | 201 (61.1) | 128 (64.6) | 9607 (65.4) |
| **Grade 1** | 40 (24.7) | 47 (28.1) | 87 (26.4) | 44 (22.2) | 3431 (23.4) |
| **Grade 2** | 50 (30.9) | 44 (26.3) | 94 (28.6) | 66 (33.3) | 4743 (32.3) |
| **Grade 3** | 13 (8.0) | 7 (4.2) | 20 (6.1) | 18 (9.1) | 1433 (9.8) |
| **Myalgia, N1** | 162 | 167 | 329 | 198 | 14687 |
| **Any** | 86 (53.1) | 82 (49.1) | 168 (51.1) | 104 (52.5) | 8529 (58.1) |
| **Grade 1** | 40 (24.7) | 47 (28.1) | 87 (26.4) | 35 (17.7) | 3242 (22.1) |
| **Grade 2** | 37 (22.8) | 30 (18.0) | 67 (20.4) | 54 (27.3) | 3966 (27.0) |
| **Grade 3** | 9 (5.6) | 5 (3.0) | 14 (4.3) | 15 (7.6) | 1321 (9.0) |
| Arthralgia, N1 | 162 | 167 | 329 | 198 | 14687 |
| Any | 66 (40.7) | 69 (41.3) | 135 (41.0) | 77 (38.9) | 6303 (42.9) |
| Grade 1 | 35 (21.6) | 43 (25.7) | 78 (23.7) | 32 (16.2) | 2809 (19.1) |
| Grade 2 | 23 (14.2) | 21 (12.6) | 44 (13.4) | 37 (18.7) | 2719 (18.5) |
| Grade 3 | 8 (4.9) | 5 (3.0) | 13 (4.0) | 8 (4.0) | 775 (5.3) |
| Nausea/Vomiting, N1 | 162 | 167 | 329 | 198 | 14687 |
| Any | 29 (17.9) | 19 (11.4) | 48 (14.6) | 41 (20.7) | 2794 (19.0) |
| Grade 1 | 25 (15.4) | 16 (9.6) | 41 (12.5) | 25 (12.6) | 2094 (14.3) |
| Grade 2 | 4 (2.5) | 3 (1.8) | 7 (2.1) | 16 (8.1) | 678 (4.6) |
| Grade 3 | 0 | 0 | 0 | 0 | 21 (0.1) |
| Grade 4 | 0 | 0 | 0 | 0 | 1 (<0.1) |
| Chills, N1 | 162 | 167 | 329 | 198 | 14687 |
| Any | 62 (38.3) | 59 (35.3) | 121 (36.8) | 78 (39.4) | 6500 (44.3) |
| Grade 1 | 32 (19.8) | 36 (21.6) | 68 (20.7) | 30 (15.2) | 2907 (19.8) |
| Grade 2 | 28 (17.3) | 23 (13.8) | 51 (15.5) | 47 (23.7) | 3402 (23.2) |
| Grade 3 | 2 (1.2) | 0 | 2 (0.6) | 1 (0.5) | 191 (1.3) |

N1 = Number of exposed subjects who submitted any data for the event. NR = not reported.
Percentages are based on the number of exposed subjects who submitted any data for the event (N1).

**Supplementary Table 3:** Solicited Adverse Reactions Within 7 Days After the Booster injection by Age Groups– Solicited Safety Set.

| Adverse Reaction<br>n (%) | ≥18 to <55 yrs |  |  | ≥55 yrs |  |  |
| --- | --- | --- | --- | --- | --- | --- |
|  | 50 µg Prime<br>N=73 | 100 µg<br>Prime<br>N=79 | Pooled 50 µg<br>+ 100 µg<br>Prime<br>N=152 | 50 µg Prime<br>N=90 | 100 µg<br>Prime<br>N=88 | Pooled 50 µg<br>+ 100 µg<br>Prime<br>N=178 |
| Solicited AR, N1 | 73 | 79 | 152 | 90 | 88 | 178 |
| Any Solicited AR | 68 (93.2) | 73 (92.4) | 141 (92.8) | 84 (93.3) | 78 (88.6) | 162 (91.0) |
| Grade 1 | 33 (45.2) | 30 (38.0) | 63 (41.4) | 32 (35.6) | 43 (48.9) | 75 (42.1) |
| Grade 2 | 24 (32.9) | 33 (41.8) | 57 (37.5) | 34 (37.8) | 26 (29.5) | 60 (33.7) |
| Grade 3 | 11 (15.1) | 9 (11.4) | 20 (13.2) | 18 (20.0) | 9 (10.2) | 27 (15.2) |
| Any Solicited Local AR, N1 | 72 | 79 | 151 | 90 | 88 | 178 |
| Any Solicited Local AR | 66 (91.7) | 69 (87.3) | 135 (89.4) | 78 (86.7) | 74 (84.1) | 152 (85.4) |
| Grade 1 | 49 (68.1) | 48 (60.8) | 97 (64.2) | 53 (58.9) | 60 (68.2) | 113 (63.5) |
| Grade 2 | 13 (18.1) | 17 (21.5) | 30 (19.9) | 20 (22.2) | 10 (11.4) | 30 (16.9) |
| Grade 3 | 4 (5.6) | 4 (5.1) | 8 (5.3) | 5 (5.6) | 4 (4.5) | 9 (5.1) |
| Local AR, Pain, N1 | 72 | 79 | 151 | 90 | 88 | 178 |
| Pain | 66 (91.7) | 68 (86.1) | 134 (88.7) | 78 (86.7) | 72 (81.8) | 150 (84.3) |
| Grade 1 | 51 (70.8) | 50 (63.3) | 101 (66.9) | 60 (66.7) | 61 (69.3) | 121 (68.0) |
| Grade 2 | 11 (15.3) | 15 (19.0) | 26 (17.2) | 15 (16.7) | 8 (9.1) | 23 (12.9) |
| Grade 3 | 4 (5.6) | 3 (3.8) | 7 (4.6) | 3 (3.3) | 3 (3.4) | 6 (3.4) |
| Erythema, N1 | 72 | 79 | 151 | 90 | 88 | 178 |
| Erythema | 4 (5.6) | 5 (6.3) | 9 (6.0) | 6 (6.7) | 3 (3.4) | 9 (5.1) |
| Grade 1 | 2 (2.8) | 2 (2.5) | 4 (2.6) | 2 (2.2) | 3 (3.4) | 5 (2.8) |
| Grade 2 | 2 (2.8) | 2 (2.5) | 4 (2.6) | 2 (2.2) | 0 | 2 (1.1) |
| Grade 3 | 0 | 1 (1.3) | 1 (0.7) | 2 (2.2) | 0 | 2 (1.1) |
| Swelling, N1 | 72 | 79 | 151 | 90 | 88 | 178 |
| Swelling | 5 (6.9) | 5 (6.3) | 10 (6.6) | 7 (7.8) | 4 (4.5) | 11 (6.2) |
| Grade 1 | 3 (4.2) | 3 (3.8) | 6 (4.0) | 1 (1.1) | 1 (1.1) | 2 (1.1) |
| Grade 2 | 2 (2.8) | 2 (2.5) | 4 (2.6) | 5 (5.6) | 2 (2.3) | 7 (3.9) |
| Grade 3 | 0 | 0 | 0 | 1 (1.1) | 1 (1.1) | 2 (1.1) |
| Axillary Swelling, N1 | 72 | 79 | 151 | 90 | 88 | 178 |
| Axillary Swelling | 17 (23.6) | 22 (27.8) | 39 (25.8) | 18 (20.0) | 12 (13.6) | 30 (16.9) |
| Grade 1 | 11 (15.3) | 18 (22.8) | 29 (19.2) | 11 (12.2) | 12 (13.6) | 23 (12.9) |
| Grade 2 | 6 (8.3) | 3 (3.8) | 9 (6.0) | 7 (7.8) | 0 | 7 (3.9) |
| Grade 3 | 0 | 1 (1.3) | 1 (0.7) | 0 | 0 | 0 |
| Any Systemic AR, N1 | 73 | 79 | 152 | 90 | 88 | 178 |
| Any Systemic AR | 55 (75.3) | 59 (74.7) | 114 (75.0) | 72 (80.0) | 67 (76.1) | 139 (78.1) |
| Grade 1 | 24 (32.9) | 23 (29.1) | 47 (30.9) | 25 (27.8) | 37 (42.0) | 62 (34.8) |
| Grade 2 | 22 (30.1) | 29 (36.7) | 51 (33.6) | 34 (37.8) | 24 (27.3) | 58 (32.6) |
| Grade 3 | 8 (11.0) | 6 (7.6) | 14 (9.2) | 13 (14.4) | 6 (6.8) | 19 (10.7) |
| Fever, N1 | 72 | 79 | 151 | 90 | 87 | 177 |
| Fever | 4 (5.6) | 6 (7.6) | 10 (6.6) | 9 (10.0) | 5 (5.7) | 14 (7.9) |
| Grade 1 | 4 (5.6) | 4 (5.1) | 8 (5.3) | 8 (8.9) | 2 (2.3) | 10 (5.6) |
| Grade 2 | 0 | 1 (1.3) | 1 (0.7) | 1 (1.1) | 2 (2.3) | 3 (1.7) |
| Grade 3 | 0 | 1 (1.3) | 1 (0.7) | 0 | 1 (1.1) | 1 (0.6) |
| Headache, N1 | 72 | 79 | 151 | 90 | 88 | 178 |
| Headache | 41 (56.9) | 45 (57.0) | 86 (57.0) | 56 (62.2) | 47 (53.4) | 103 (57.9) |
| Grade 1 | 23 (31.9) | 28 (35.4) | 51 (33.8) | 34 (37.8) | 33 (37.5) | 67 (37.6) |
| Grade 2 | 16 (22.2) | 16 (20.3) | 32 (21.2) | 18 (20.0) | 13 (14.8) | 31 (17.4) |
| Grade 3 | 2 (2.8) | 1 (1.3) | 3 (2.0) | 4 (4.4) | 1 (1.1) | 5 (2.8) |
| Fatigue, N1 | 72 | 79 | 151 | 90 | 88 | 178 |
| Fatigue | 44 (61.1) | 46 (58.2) | 90 (59.6) | 59 (65.6) | 52 (59.1) | 111 (62.4) |
| Grade 1 | 18 (25.0) | 18 (22.8) | 36 (23.8) | 22 (24.4) | 29 (33.0) | 51 (28.7) |
| Grade 2 | 22 (30.6) | 25 (31.6) | 47 (31.1) | 28 (31.1) | 19 (21.6) | 47 (26.4) |
| Grade 3 | 4 (5.6) | 3 (3.8) | 7 (4.6) | 9 (10.0) | 4 (4.5) | 13 (7.3) |
| Myalgia, N1 | 72 | 79 | 151 | 90 | 88 | 178 |
| Myalgia | 37 (51.4) | 37 (46.8) | 74 (49.0) | 49 (54.4) | 45 (51.1) | 94 (52.8) |
| Grade 1 | 20 (27.8) | 19 (24.1) | 39 (25.8) | 20 (22.2) | 28 (31.8) | 48 (27.0) |
| Grade 2 | 13 (18.1) | 15 (19.0) | 28 (18.5) | 24 (26.7) | 15 (17.0) | 39 (21.9) |
| Grade 3 | 4 (5.6) | 3 (3.8) | 7 (4.6) | 5 (5.6) | 2 (2.3) | 7 (3.9) |
| Arthralgia, N1 | 72 | 79 | 151 | 90 | 88 | 178 |
| Arthralgia | 24 (33.3) | 34 (43.0) | 58 (38.4) | 42 (46.7) | 35 (39.8) | 77 (43.3) |
| Grade 1 | 13 (18.1) | 20 (25.3) | 33 (21.9) | 22 (24.4) | 23 (26.1) | 45 (25.3) |
| Grade 2 | 7 (9.7) | 12 (15.2) | 19 (12.6) | 16 (17.8) | 9 (10.2) | 25 (14.0) |
| Grade 3 | 4 (5.6) | 2 (2.5) | 6 (4.0) | 4 (4.4) | 3 (3.4) | 7 (3.9) |
| Nausea/<br>vomiting, N1 | 72 | 79 | 151 | 90 | 88 | 178 |
| Nausea /<br>vomiting | 17 (23.6) | 12 (15.2) | 29 (19.2) | 12 (13.3) | 7 (8.0) | 19 (10.7) |
| Grade 1 | 15 (20.8) | 12 (15.2) | 27 (17.9) | 10 (11.1) | 4 (4.5) | 14 (7.9) |
| Grade 2 | 2 (2.8) | 0 | 2 (1.3) | 2 (2.2) | 3 (3.4) | 5 (2.8) |
| Grade 3 | 0 | 0 | 0 | 0 | 0 | 0 |
| Chills, N1 | 72 | 79 | 151 | 90 | 88 | 178 |
| Chills | 28 (38.9) | 30 (38.0) | 58 (38.4) | 34 (37.8) | 29 (33.0) | 63 (35.4) |
| Grade 1 | 15 (20.8) | 20 (25.3) | 35 (23.2) | 17 (18.9) | 16 (18.2) | 33 (18.5) |
| Grade 2 | 12 (16.7) | 10 (12.7) | 22 (14.6) | 16 (17.8) | 13 (14.8) | 29 (16.3) |
| Grade 3 | 1 (1.4) | 0 | 1 (0.7) | 1 (1.1) | 0 | 1 (0.6) |

N1=Number of exposed participants who submitted any data for the event.
Percentages are based on the number of exposed participants who submitted any data for the event (N1).

**Supplementary Table 4:** Neutralizing Antibody Titers (Pseudovirus ID50 versus D614G) after the Primary Series and a Booster Injection of 50 µg of mRNA-1273 (Per-protocol Set)

|  | 50 µg Prime<br>N=146 | 100 µg Prime<br>N=149 | 50 µg and 100<br>µg Prime<br>N=295 |
| --- | --- | --- | --- |
| Baseline (Day 1), n* | 146 | 148 | 294 |
| GMT | 9.4 | 9.3 | 9.3 |
| 95% CI | 9.2, 9.5 | NE | 9.2, 42.7 |
| Day 29 (28 Days after 1st injection), n | 144 | 146 | 290 |
| GMT | 27.3 | 36.4 | 31.6 |
| 95% CI | 23.3, 31.8 | 30.8, 43.1 | 28.1, 35.4 |
| Day 57 (28 Days after 2<sup>nd</sup> injection), n | 143 | 146 | 289 |
| GMT | 629.2 | 1268.0 | 896.5 |
| 95% CI | 549.3, 720.8 | 1087.9, 1477.8 | 803.4, 1000.4 |
| Open-label, Day 1 (OL-D1; Pre-Booster), n* | 145 | 149 | 294 |
| GMT | 104.7 | 150.2 | 125.7 |
| 95% CI<sup>†</sup> | 88.3, 124.1 | 125.7, 179.5 | 111.0, 142.3 |
| Open-label Day 29 (OL-D29; 28 Days After Booster), n<sup>‡</sup> | 146 | 149 | 295 |
| GMT | 1834.3 | 1951.7 | 1892.7 |
| 95% CI | 1600.2, 2102.6 | 1729.6, 2202.4 | 1728.8, 2072.2 |
| Comparison of OL-D29 after the booster to Day 57, n | 143 | 146 | 289 |
| GM Fold Rise | 2.9 | 1.5 | 2.1 |
| 95% CI | 2.6, 3.4 | 1.3, 1.8 | 1.9, 2.3 |

Antibody values reported as below the lower limit of quantification (LLOQ; 18.5) were replaced by 0.5 x LLOQ. Values that were greater than the upper limit of quantification (ULOQ; 45118) were converted to the ULOQ if actual values were not available.
\*n=Number of subjects with non-missing baseline results.
<sup>†</sup>95% Confidence Intervals were calculated based on the t-distribution of the log-transformed values or the difference in the log-transformed values for GMT and GMFR, respectively, then back transformed to the original scale.
<sup>‡</sup> Number of subjects in the Per-Protocol Set with non-missing data at the corresponding visit.

**Supplementary Table 5:** Comparison of Antibody Responses Versus D614G After Boosting with Those After Completion of Primary Series.

|  | Boosting with 50 µg of mRNA-1273 after primary series* [Phase 2 Part B] | Primary Series of Two Injections of 100 µg of mRNA-1273 [Phase 3 COVE random sub-cohort] |
| --- | --- | --- |
| **Pseudovirus Neutralizing Antibody (ID50)** |  |  |
| Baseline, n | 294 | 1052 |
| Baseline, GMT (95% CI) | 125.7 (111.0, 142.3) | 9.6 (9.4, 9.9) |
| GMT at 28 Days after boosting or at 28 Days after second injection of mRNA-1273 during primary series (95% CI) | 1892.7 (1728.8, 2072.2) | 1081.1 (1019.8, 1146.1) |
| Seroresponse Rate†, n | 294 | 1050 |
| % (95% CI) | 90.1 (86.1, 93.3) | 98.4 (97.4, 99.1) |
| Difference in seroresponse rates†, % (95% CI) | -8.2 (-12.2, -5.2) |  |
| **Anti-Spike ELISA (VAC65)** |  |  |
| Baseline, n | 290 | 1052 |
| Baseline GMT (95% CI) | 97.4 (89.4, 106.1) | 0.7 (0.7, 0.8) |
| GMT at 28 Days after boosting or at 28 Days after second injection of mRNA-1273 during primary series (95% CI) | 1074.7 (1024.1, 1127.8) | 694.8 (664.8, 726.1) |
| Seroresponse Rate, % (95% CI) | 94.7 (91.4, 97.0) | 99.6 (99.0, 99.9) |
| Difference in seroresponse rates, % (95% CI)† | -4.9 (-8.2, -2.8) |  |
| **Anti-Spike IgG Antibody by MSD MULTIPLEX** |  |  |
| Baseline, n | 290 | 1046 |
| Baseline, GMT (95% CI) | 34,827.3 (31,546.4, 38,449.5) | 114.8 (110.8, 119.0) |
| 28 Days after boosting or at 28 Days after second injection of mRNA-1273 during primary series, n | 289 | 1035 |
| GMT (95% CI) | 569,716.1 (536,434.6, 605,062.5) | 316,448.3 (300,071.4, 333,719.0) |
| Seroresponse Rate, n | 284 | 1027 |
| % (95% CI) | 98.2 (95.9, 99.4) | 99.4 (98.7, 99.8) |
| Difference in seroresponse rates, % (95% CI)† | -1.2 (-3.5, 0.0) |  |

\* Overall: Combined 50 µg and 100 µg prime groups
† Seroresponse at participant level was defined as a change of titer from below the lower limit of quantification (LLOQ) to equal to or above 4 times the LLOQ, or a 4-times or higher ratio in participants with titers above LLOQ. The difference in seroresponse rates was Boosting minus Primary Series. For participants who received the primary series, seroresponse was defined based on the fold-rise from baseline titer (prior to first dose of primary dose). For participants who received a booster vaccination, seroresponse was defined based on the fold-rise from pre-booster titer (at least 6 months after completion of the primary series).
GMT: Observed

**Supplementary Table 6:** Spike Binding IgG Antibody versus D614G by ELISA (VAC65) After the Primary Series and a 50 µg Booster Injection – Per-protocol Immunogenicity Subset.

|  | 50 µg Prime | 100 µg Prime | 50 µg and 100 µg Prime |
| --- | --- | --- | --- |
| Baseline (Day 1), n* | 146 | 148 | 294 |
| GMT | 0.7 | 0.67 | 0.7 |
| 95% CI | 0.6, 0.8 | 0.6, 0.7 | 0.6, 0.7 |
| 28 days after 1st injection, n | 145 | 146 | 291 |
| GMT | 59.1 | 79.3 | 68.5 |
| 95% CI | 50.8, 68.6 | 68.0, 92.5 | 61.4, 76.3 |
| 28 days after 2nd injection, n | 146 | 147 | 293 |
| GMT | 514.4 | 640.7 | 574.3 |
| 95% CI | 464.4, 569.7 | 575.5, 713.2 | 533.0, 618.8 |
| Pre-booster, n | 144 | 146 | 290 |
| GMT | 86.4 | 109.6 | 97.4 |
| 95% CI | 76.9, 97.0 | 96.8, 124.2 | 89.4, 106.1 |
| Day 28 After Booster, n | 142 | 149 | 289 |
| GMT | 1068.8 | 1080.4 | 1074.7 |
| 95% CI | 991.6, 1152.1 | 1015.5, 1149.4 | 1024.1, 1127.8 |
| Comparison of 28 days after booster to 28 days after primary series, n | 142 | 145 | 287 |
| GM Fold Rise | 2.1 | 1.7 | 1.9 |
| 95% CI | 1.9, 2.2 | 1.5, 1.9 | 1.7, 2.0 |

\*n = Number of participants in the Per-Protocol Set with non-missing data at baseline and the corresponding visit.
Antibody values reported as below the lower limit of quantification (LLOQ) were replaced by 0.5 x LLOQ. Values that were greater than the upper limit of quantification (ULOQ) were converted to the ULOQ if actual values are not available. Percentages are based on the number of participants in the Per-Protocol Set with non-missing data at baseline and the corresponding visit. 95% Confidence Intervals were calculated based on the t-distribution of the log-transformed values or the difference in the log-transformed values for GMT and GMFR, respectively, then back transformed to the original scale for presentation.

**Supplementary Table 7:** Unsolicited Treatment-emergent Adverse Events (TEAEs) up to 28 days after Booster Injection.

|  | **50 µg**<br>**Prime,**<br>**Booster**<br>**(N=173)**<br>**n (%)** | **100 µg**<br>**Prime,**<br>**Booster**<br>**(N=171)**<br>**n (%)** | **50 µg and 100**<br>**µg Prime,**<br>**Booster**<br>**(N=344)**<br>**n (%)** |
| --- | --- | --- | --- |
| Unsolicited TEAEs regardless of relationship to study vaccination |  |  |  |
| All | 17 (9.8) | 22 (12.9) | 39 (11.3) |
| Serious | 0 | 0 | 0 |
| Fatal | 0 | 0 | 0 |
| Medically-attended | 8 (4.6) | 12 (7.0) | 20 (5.8) |
| Leading to study discontinuation | 0 | 0 | 0 |
| Severe | 0 | 0 | 0 |
| Unsolicited TEAEs related to study vaccination |  |  |  |
| All | 6 (3.5) | 7 (4.1) | 13 (3.8) |
| Serious | 0 | 0 | 0 |
| Fatal | 0 | 0 | 0 |
| Medically-attended | 0 | 2 (1.2) | 2 (0.6) |
| Leading to study discontinuation | 0 | 0 | 0 |
| Severe | 0 | 0 | 0 |

**Supplementary Table 8:** Neutralizing Antibody Titers (Pseudovirus ID50 versus the Delta Variant) after the Primary Series and a Booster Injection of 50 µg of mRNA-1273 (Per-protocol Set)

|  | 50 µg Prime<br>N=146 | 100 µg Prime<br>N=149 | 50 µg and 100<br>µg Prime<br>N=295 |
| --- | --- | --- | --- |
| OL-Day 1 (Pre-booster), n* | 144 | 149 | 293 |
| GMT | 37.1 | 47.9 | 42.3 |
| 95% CI | 31.3, 44.2 | 39.7, 57.8 | 37.2, 48.0 |
| OL-Day 29 (28 Days after booster), n<sup>‡</sup> | 146 | 149 | 295 |
| GMT | 779.5 | 827.8 | 803.5 |
| 95% CI | 670.1, 906.8 | 738.5, 927.9 | 731.4, 882.7 |
| Comparison of OL-D29 to OL-D1 |  |  |  |
| GM Fold Rise | 20.9 | 17.3 | 19.0 |
| 95% CI | 17.5, 24.9 | 14.4, 20.8 | 16.7, 21.5 |

\*n=Number of participants with non-missing results at pre-booster.
<sup>†</sup>95% Confidence Intervals were calculated based on the t-distribution of the log-transformed values or the difference in the log-transformed values for GMT and GMFR, respectively, then back transformed to the original scale.
<sup>‡</sup> Number of participants in the Per-Protocol Set with non-missing data at the corresponding visit.

## REFERENCES

1. FDA. FDA Authorizes Booster Dose of Pfizer-BioNTech COVID-19 Vaccine for Certain Populations, <https://www.fda.gov/news-events/press-announcements/fda-authorizes-booster-dose-pfizer-biontech-covid-19-vaccine-certain-populations> (2021).

2. Pormohammad, A. et al. Efficacy and Safety of COVID-19 Vaccines: A Systematic Review and Meta-Analysis of Randomized Clinical Trials. Vaccines (Basel) 9, doi:10.3390/vaccines9050467 (2021).

3. H. Ritchie, E. M, L. Rodés-Guirao, C. Appel, C. Giattino, E. Ortiz-Ospina, J. Hasell, B. Macdonald, D. Beltekian, M. Roser. Coronavirus Pandemic (COVID-19), <https://ourworldindata.org/coronavirus> (2021).

4. FDA. FDA Approves First COVID-19 Vaccine, <https://www.fda.gov/news-events/press-announcements/fda-approves-first-covid-19-vaccine> (2021).

5. Baden, L. R. et al. Efficacy and Safety of the mRNA-1273 SARS-CoV-2 Vaccine. N Engl J Med 384, 403–416, doi:10.1056/NEJMoa2035389 (2021).

6. Polack, F. P. et al. Safety and Efficacy of the BNT162b2 mRNA Covid-19 Vaccine. N Engl J Med 383, 2603–2615, doi:10.1056/NEJMoa2034577 (2020).

7. Sadoff, J. et al. Interim Results of a Phase 1-2a Trial of Ad26.COV2.S Covid-19 Vaccine. N Engl J Med 384, 1824–1835, doi:10.1056/NEJMoa2034201 (2021).

8. Ali, K. et al. Evaluation of mRNA-1273 SARS-CoV-2 Vaccine in Adolescents. N Engl J Med, doi:10.1056/NEJMoa2109522 (2021).

9. Jackson, L. A. et al. An mRNA Vaccine against SARS-CoV-2 -Preliminary Report. N Engl J Med 383, 1920–1931, doi:10.1056/NEJMoa2022483 (2020).

10. Anderson, E. J. et al. Safety and Immunogenicity of SARS-CoV-2 mRNA-1273 Vaccine in Older Adults. N Engl J Med 383, 2427–2438, doi:10.1056/NEJMoa2028436 (2020).

11. Chu, L. et al. A preliminary report of a randomized controlled phase 2 trial of the safety and immunogenicity of mRNA-1273 SARS-CoV-2 vaccine. Vaccine 39, 2791–2799, doi:10.1016/j.vaccine.2021.02.007 (2021).

12. FDA. Moderna COVID-19 Vaccine, <https://www.fda.gov/emergency-preparedness-and-response/coronavirus-disease-2019-covid-19/moderna-covid-19-vaccine> (2021).

13. El Sahly, H. M. et al. Efficacy of the mRNA-1273 SARS-CoV-2 Vaccine at Completion of Blinded Phase. N Engl J Med, doi:10.1056/NEJMoa2113017 (2021).

14. CDC. SARS-CoV-2 Variant Classifications and Definitions, <https://www.cdc.gov/coronavirus/2019-ncov/cases-updates/variant-surveillance/variant-info.html> (2021).

15. CDC. COVID Data Tracker: Variant Proportions, <https://covid.cdc.gov/covid-data-tracker/#variant-proportions> (2021).

16. GISAID. GISAID: Overview of Variants in Countries, <https://covariants.org/per-country> (2021).

17. Pegu, A. et al. Durability of mRNA-1273-induced antibodies against SARS-CoV-2 variants. Science, doi:10.1126/science.abj4176 (2021).

18. Corbett, K. S. et al. Immune correlates of protection by mRNA-1273 vaccine against SARS-CoV-2 in nonhuman primates. *Science*, eabj0299, doi:10.1126/science.abj0299 (2021).

19. Liu, Y. et al. Neutralizing Activity of BNT162b2-Elicited Serum - Preliminary Report. N Engl J Med, doi:10.1056/NEJMc2102017 (2021).

20. Lopez Bernal, J. et al. Effectiveness of Covid-19 Vaccines against the B.1.617.2 (Delta) Variant. N Engl J Med 385, 585–594, doi:10.1056/NEJMoa2108891 (2021).

21. Abu-Raddad, L. J., Chemaitelly, H., Butt, A. A. & National Study Group for, C.-V. Effectiveness of the BNT162b2 Covid-19 Vaccine against the B.1.1.7 and B.1.351 Variants. N Engl J Med 385, 187–189, doi:10.1056/NEJMc2104974 (2021).

22. Wu, K. et al. Serum Neutralizing Activity Elicited by mRNA-1273 Vaccine - Preliminary Report. N Engl J Med, doi:10.1056/NEJMc2102179 (2021).

23. Choi, A. et al. Safety and immunogenicity of SARS-CoV-2 variant mRNA vaccine boosters in healthy adults: an interim analysis. Nature Medicine, doi:https://doi.org/10.1038/s41591-021-01527-y (2021).

24. Puranik, A. et al. Comparison of two highly-effective mRNA vaccines for COVID-19 during periods of Alpha and Delta variant prevalence. medRxiv, doi:10.1101/2021.08.06.21261707 (2021).

25. Nanduri, S. et al. Effectiveness of Pfizer-BioNTech and Moderna Vaccines in Preventing SARS-CoV-2 Infection Among Nursing Home Residents Before and During Widespread Circulation of the SARS-CoV-2 B.1.617.2 (Delta) Variant -National Healthcare Safety Network, March 1-August 1, 2021. MMWR Morb Mortal Wkly Rep 70, 1163–1166, doi:10.15585/mmwr.mm7034e3 (2021).

26. Tang, P. et al. BNT162b2 and mRNA-1273 COVID-19 vaccine effectiveness against the Delta (B.1.617.2) variant in Qatar. MedRxiv, doi:https://doi.org/10.1101/2021.08.11.21261885 (2021).

27. Fowlkes, A. et al. Effectiveness of COVID-19 Vaccines in Preventing SARS-CoV-2 Infection Among Frontline Workers Before and During B.1.617.2 (Delta) Variant Predominance - Eight U.S. Locations, December 2020-August 2021. MMWR Morb Mortal Wkly Rep 70, 1167–1169, doi:10.15585/mmwr.mm7034e4 (2021).

28. Chu, L. et al. A preliminary report of a randomized controlled phase 2 trial of the safety and immunogenicity of mRNA-1273 SARS-CoV-2 vaccine. Vaccine 39, 2791–2799, doi:10.1016/j.vaccine.2021.02.007 (2021).

29. Shen, X. et al. SARS-CoV-2 variant B.1.1.7 is susceptible to neutralizing antibodies elicited by ancestral spike vaccines. Cell Host Microbe 29, 529–539 e523, doi:10.1016/j.chom.2021.03.002 (2021).

30. Akkaya, M., Kwak, K. & Pierce, S. K. B cell memory: building two walls of protection against pathogens. Nat Rev Immunol 20, 229–238, doi:10.1038/s41577-019-0244-2 (2020).

31. Dan, J. M. et al. Immunological memory to SARS-CoV-2 assessed for up to 8 months after infection. Science 371, doi:10.1126/science.abf4063 (2021).

32. Goel, R. R. et al. mRNA Vaccination Induces Durable Immune Memory to SARS-CoV-2 with Continued Evolution to Variants of Concern. bioRxiv, doi:10.1101/2021.08.23.457229 (2021).

33. Sallusto, F., Lanzavecchia, A., Araki, K. & Ahmed, R. From vaccines to memory and back. Immunity 33, 451–463, doi:10.1016/j.immuni.2010.10.008 (2010).

34. Tenforde, M. W. et al. Sustained Effectiveness of Pfizer-BioNTech and Moderna Vaccines Against COVID-19 Associated Hospitalizations Among Adults - United States, March-July 2021. MMWR Morb Mortal Wkly Rep 70, 1156–1162, doi:10.15585/mmwr.mm7034e2 (2021).

35. Rosenberg, E. S. et al. New COVID-19 Cases and Hospitalizations Among Adults, by Vaccination Status - New York, May 3-July 25, 2021. MMWR Morb Mortal Wkly Rep 70, 1150–1155, doi:10.15585/mmwr.mm7034e1 (2021).

36. Self, W. H. et al. Comparative Effectiveness of Moderna, Pfizer-BioNTech, and Janssen (Johnson & Johnson) Vaccines in Preventing COVID-19 Hospitalizations Among Adults Without Immunocompromising Conditions — United States, March–August 2021. Morbidity and Mortality Weekly Report 70 https://www.cdc.gov/mmwr/volumes/70/wr/mm7038e1.htm?s_cid=mm7038e1_w (2021).

37. Gilbert, P. B. et al. Immune Correlates Analysis of the mRNA-1273 COVID-19 Vaccine Efficacy Trial MedRxiv, doi: https://doi.org/10.1101/2021.08.09.21261290 (2021).

